# Associations of plasma omega-6 and omega-3 fatty acids with overall and 19 site-specific cancers: a population-based cohort study in UK Biobank

**DOI:** 10.1101/2024.01.21.24301568

**Authors:** Yuchen Zhang, Yitang Sun, Suhang Song, Nikhil K. Khankari, J. Thomas Brenna, Ye Shen, Kaixiong Ye

**Affiliations:** Department of Epidemiology and Biostatistics, College of Public Health, University of Georgia, Athens, Georgia, US; Department of Genetics, University of Georgia, Athens, Georgia, US; Department of Health Policy and Management, College of Public Health, University of Georgia, Athens, Georgia, US; Division of Genetic Medicine, Vanderbilt University Medical Center, Nashville, Tennessee, US; Vanderbilt Genetics Institute, Vanderbilt University Medical Center, Nashville, Tennessee, US; Division of Nutritional Sciences, Cornell University, Ithaca, NY, US; Dell Pediatric Research Institute and the Depts of Pediatrics, of Nutrition, and of Chemistry, University of Texas at Austin, Austin, TX, US; Institute of Bioinformatics, University of Georgia, Athens, Georgia, US

**Keywords:** Polyunsaturated fatty acids, Omega-6 fatty acids, Omega-3 fatty acids, Cancer incidence, Prospective cohort study

## Abstract

**Background:** Previous epidemiological studies of the associations between polyunsaturated fatty acids (PUFAs) and cancer incidence have been inconsistent. We investigated the associations of plasma omega-3 and omega-6 PUFAs with the incidence of overall and 19 site-specific cancers in a large prospective cohort.

**Methods:** 253,138 eligible UK Biobank participants were included in our study. With a mean follow-up of 12.9 years, 29,838 participants were diagnosed with cancer. The plasma levels of omega-3 and omega-6 PUFAs were expressed as percentages of total fatty acids (omega-3% and omega-6%).

**Results:** In our main models, both omega-6% and omega-3% were inversely associated with overall cancer incidence (HR per SD = 0.98, 95% CI = 0.96-0.99; HR per SD = 0.99, 95% CI = 0.97-1.00; respectively). Of the 19 site-specific cancers available, 14 were associated with omega-6% and five with omega-3%, all indicating inverse associations, with the exception that prostate cancer was positively associated with omega-3% (HR per SD = 1.03, 95% CI = 1.01 – 1.05).

**Conclusions:** Our population-based cohort study in UK Biobank indicates small inverse associations of plasma omega-6 and omega-3 PUFAs with the incidence of overall and most site-specific cancers, although there are notable exceptions, such as prostate cancer.

## Background

Cancer is a leading cause of morbidity and mortality worldwide, with an estimated 18.1 million cancer cases globally in 2020. Breast, lung, and colorectal cancer account for over 30% of the total annual incidence [1]. Polyunsaturated fatty acids (PUFAs) have been postulated to influence cancer incidence and survival [2–4]. Potential mechanisms of PUFAs in cancer etiology include serving as precursors to lipid mediators regulating metabolic pathways and inflammatory responses [5], and altering membrane composition that could affect cell signaling pathways [6].

Despite extensive interest and research, the links between PUFAs and cancer remain inconclusive. An umbrella review of meta-analyses of observational studies of cancer incidence concluded that there was no convincing evidence regarding the effects of omega-3 PUFAs on the risk of any cancer, and that there was only weak evidence supporting inverse associations of omega-3 intake with liver, breast, and brain cancers [3]. A meta-analysis of observational studies of cancer survival found that the intake of fish or marine omega-3 PUFAs, but not total omega-3 PUFAs, was associated with lower mortality in cancer patients [7]. A meta-analysis of randomized trials showed that increasing marine omega-3 PUFAs had little or no effects on overall cancer diagnosis or cancer death, while the effects of increasing omega-6 PUFAs were unclear because the evidence was of very low quality [2]. These systemic reviews showcase the limitations of existing studies, which include large between-study heterogeneity, small study bias, insufficient case numbers, and short follow-up time. Moreover, most studies relied on self-reported fish oil supplementation or estimated dietary intake, which may suffer from recall errors, outdated food databases, and measurement inaccuracy [8]. Circulating biomarkers provide more objective measures of omega-3 and omega-6 PUFA status and are reflective of dietary intakes [9]. Indeed, a meta-analysis of prospective studies found that the blood level of omega-6 PUFAs, but not their intake, was inversely associated with overall cancer risk [10]. Similarly, another meta-analysis showed that the blood level of omega-3 PUFAs, but not their intake, was associated with a lower colorectal cancer risk [11]. Addressing the limitations of current studies and examining objective blood levels of PUFAs may offer clarity into the roles of omega-3 and omega-6 PUFAs in cancer risk.

UK Biobank is a large population-based prospective cohort that has followed over 500,000 participants since 2006 [12]. It is a large homogeneous cohort with a long follow-up time, offering an unprecedented opportunity to examine the effects of PUFAs on overall cancer and a comprehensive range of site-specific cancers. A few early studies have revealed that fish oil supplementation or dietary omega-3 PUFA intake was associated with lower incidence of colon cancer, lung cancer, or liver cancer [13–15]. Recently, UK Biobank obtained metabolomic measurements of baseline plasma samples for about 60% of the participants, a random subset of the full cohort [16]. Leveraging this valuable dataset, we previously showed that circulating levels of omega-3 and omega-6 PUFAs were both inversely associated with overall cancer mortality [17]. In this study, we aim to examine the associations of circulating omega-3 and omega-6 PUFAs, as well as their ratio (i.e., omega-6/omega-3), with the incidence of overall and 19 site-specific cancers in UK Biobank.

## Methods

### Study population

Between 2006 and 2010, UK Biobank recruited over half a million participants, aged 37-73, in 22 assessment centers across England, Wales and Scotland. During the baseline assessment visit, a wide variety of sociodemographic, lifestyle, and health-related data were acquired through self-administered touch-screen questionnaires, concise computer-assisted interviews, and physical and functional measures. Blood, urine, and saliva samples were also collected. Of the 502,366 participants, those who had cancer diagnoses at baseline (n=37,737, excluding nonmelanoma skin cancer with an ICD-10 code of C44), those who had withdrawn from UK Biobank (n=1,227), and those with missing data on the plasma polyunsaturated fatty acids (n=210,264) were excluded from our study. A total of 253,138 eligible participants were eventually included.

### Ascertainment of exposures

The absolute concentrations of plasma polyunsaturated fatty acids (PUFAs) were assessed using nuclear magnetic resonance (NMR) in plasma samples obtained at the baseline visit from 2007 to 2010, and the corresponding percentages of total fatty acids were calculated [12, 16]. The omega-3 fatty acids to total fatty acids percentage (omega-3%) and the omega-6 fatty acids to total fatty acids percentage (omega-6%) were the primary exposures of interest in this study. In addition, we conducted analyses on the ratio of plasma omega-6/omega-3 PUFAs, docosahexaenoic acid to total fatty acids percentage (DHA%), and linoleic acid to total fatty acids percentage (LA%). No other individual PUFAs, except DHA and LA, were measured by the NMR metabolomic platform.

### Ascertainment of outcomes

The primary outcomes were the first incidence of overall and 19 site-specific cancers based on diagnostic records in cancer registers ascertained from National Health Service (NHS) central registers [12]. At the time of our analysis (15 August 2023), we had access to the most current health outcomes dataset (Version: July 2023), which contained cancer incidence records up to 19 December 2022. Consequently, follow-up time was calculated from the recruitment date until the aforementioned date, any cancer diagnosis or death, whichever came first. The incidence of cancer was coded according to the World Health Organization’s International Statistical Classification of Diseases (ICD)-9 or ICD-10 codes. Participants who had cancer at baseline (excluding nonmelanoma skin cancer) were excluded. ICD-9 codes were only used for pre-existing cancer and thus excluded. New cancer incidence was defined based on ICD-10 codes for overall cancer (C00-C97, excluding nonmelanoma skin cancer, C44) and the following 19 site-specific cancers: head and neck (C00-C14), esophagus (C15), stomach (C16), colon (C18), rectum (C19-C20), hepatobiliary tract (C22-C24), pancreas (C25), lung (C33-C34), malignant melanoma (C43), connective soft tissue (C49), breast (C50), uterus (C54-C55), ovary (C56), prostate (C61), kidney (C64-C65), bladder (C66-C67), brain (C70-C72), thyroid (C73), and lymphoid and hematopoietic tissues (C81-C96).

### Covariates

The initial questionnaire covered a comprehensive range of potential confounding factors: demographic characteristics (e.g., age, gender, ethnicity); socioeconomic status, as measured by the Townsend Deprivation Index (TDI); lifestyle behaviors (e.g., alcohol consumption, smoking status, body mass index (BMI), and physical activity); and history or family history of diseases (e.g., diabetes, gastroesophageal reflux disease and family history of cancer). Body mass index (BMI) was calculated from weight and height expressed in kg/m^2^. Waist circumference and hip circumference were recorded at a central registry, and we calculated the corresponding waist-hip ratio (waist circumference divided by hip circumference). The TDI, employed as a measure of socioeconomic deprivation, was directly obtained from the UK Biobank database, with a higher score indicating a higher level of socioeconomic deprivation.

### Statistical analyses

We began by summarizing and comparing participant characteristics based on the quintiles of the plasma omega-6% and omega-3% at baseline using descriptive statistics. To assess the differences in demographic features across these quintiles, we employed Pearson’s Chi-squared test for categorical variables and the ANOVA test for continuous variables.

To explore the associations with cancer incidence for plasma omega-6%, omega-3%, and their ratio, we utilized multivariable Cox proportional hazards regression models to estimate hazard ratios (HRs) along with their corresponding 95% confidence intervals (CIs). We developed three distinct models, namely, the simply adjusted model, the main model, and the additionally adjusted model. Within the simply adjusted model, age and sex were designed as stratification variables owing to their violation of the assumptions inherent to the proportional hazards model. The main model was additionally adjusted for ethnicity (classified into White, Black, Asian, Others), TDI (continuous), assessment center (categorical), BMI (kg/m2; continuous), smoking status (categorized as never, previous, current), alcohol intake status (categorized as never, previous, current), and physical activity (classified as low, moderate, high). In addition to investigating the overall cancer, we also performed separate analyses for each site-specific cancer. The analysis of prostate cancer was restricted to the male sample, whereas the investigation of breast cancer, ovarian cancer, and uterine cancer was limited to the female sample. Furthermore, to adjust for additional possible confounding variables, we incorporated additional covariates into the analysis for certain cancer types (i.e., additionally adjusted models), guided by previous literature and biological plausibility [15]. More details can be found in Table S1.

Our analysis treated the exposures of interest both in continuous (standardized to a mean of 0 and standard deviation of 1) and categorical (in quintiles) terms. When conducting trend tests, we used the median value of each quintile as a continuous variable within the models. Given 19 distinct cancer subtypes, we adopted the False Discovery Rate (FDR) approach to address the issue of increasing false positives arising from multiple testing and reported the adjusted p-values for simply adjusted models and main models. We did not perform multiple testing correction for the additionally adjusted models because they were for the purpose of sensitivity analysis and were only performed for 10 site-specific cancers with site-specific covariates. We also evaluated potential nonlinear dose-response using a semi-parametric approach through the utilization of restricted cubic splines [18] (4 knots were used in regression splines). We considered there was evidence supporting the presence of an association between a PUFA exposure and a cancer outcome if the continuous exposure analysis or the trend across quintiles analysis was statistically significant in the main models or in the additionally adjusted models, if applicable. In addition to the two above-described analyses, we assessed if there were any differences among the HRs across the five quintiles by applying likelihood ratio tests.

In secondary analyses aiming at investigating potential variations in associations within distinct population subgroups, we replicated the aforementioned analyses for overall cancer while stratifying the data by the following factors: age (< vs. ≥ the median age of 58 years), sex (male vs. female), TDI (< vs. ≥ the population median of –2), BMI (< vs. ≥ 25), current smoking status (yes vs. no), current alcohol consumption status (yes vs. no) and level of physical activity (low and moderate vs. high). The exposures of interest (omega-6%, omega-3%, and their ratio) were categorized in quintiles. For each stratification variable, we conducted a likelihood ratio test to obtain the associated p-value for interaction. In the case of continuous stratification variables (i.e., age, TDI, and BMI), we calculated interaction p-values based on a one-unit alteration of the respective stratification variables.

Furthermore, we carried out a series of sensitivity analyses. First, to assess whether the association of plasma omega-6% with overall cancer risk would be altered by omega-3% or vice versa, we replicated the main analysis for overall cancer while involving both omega-6% and omega-3% as variables in the model. The correlation between omega-3% and omega-6% was assessed by the Pearson correlation. Second, to explore the effects of individual fatty acids, DHA and LA, on cancer incidence, we repeated the main analysis on DHA% and LA%. Third, to investigate the potential impact of reverse causation on the observed associations, individuals who experienced outcomes within the first year or the first three years of the follow-up period were excluded from the analysis. Last, to evaluate the representativeness of the study participants, we conducted a comparative analysis of baseline characteristics between those individuals with exposure information and those without it. All p-values were assessed using a two-sided approach. Statistical significance was defined as a p-value less than 0.05 or a 95% confidence interval that did not include the value 1.0 for the corresponding HRs. We conducted all analyses using R (version: 4.0.3).

## Results

### Baseline characteristics

Within our analytical cohort of 253,138 participants, spanning an average follow-up period of 12.9 years, a total of 29,838 individuals were diagnosed with cancer during follow-up. The baseline characteristics of all participants distributed across quintiles of plasma omega-6% and omega-3% were summarized in Table 1 and Table S2, respectively. On average, study participants were approximately 56 years old, with 90% of them identifying as White. Those in the higher quintiles of plasma omega-6% tended to be younger, female, with lower BMI and more physically active, and were less likely to smoke or drink alcohol.

**Table 1.** Baseline characteristics of included participants by quintiles of the plasma omega-6% (n = 253,138)

| Characteristics<sup>a</sup> | Omega-6% quintiles |  |  |  |  | p-value |
| --- | --- | --- | --- | --- | --- | --- |
|  | 1 (median = 32.9)<br>(n = 50,628) | 2 (median = 36.4)<br>(n = 50,628) | 3 (median = 38.4)<br>(n = 50,628) | 4 (median = 40.0)<br>(n = 50,627) | 5 (median = 42.1)<br>(n = 50,627) |  |
| **Age** (years) | 57.6 (7.7) | 57.7 (7.8) | 57.1 (7.9) | 55.9 (8.1) | 53.6 (8.2) | <0.001<sup>a</sup> |
| **Gender** (male%) | 61.8 | 49.8 | 42.9 | 39.6 | 41.9 | <0.001<sup>b</sup> |
| **Ethnicity**(n%) |  |  |  |  |  |  |
| White | 46,982 (93.2%) | 46,868 (92.9%) | 46,597 (92.4%) | 45,918 (91.1%) | 43,857 (87.2%) | <0.001<sup>b</sup> |
| Black | 196 (0.4%) | 229 (0.5%) | 237 (0.5%) | 256 (0.5%) | 459 (0.9%) |  |
| Asian | 1,387 (2.8%) | 1,514 (3.0%) | 1,663 (3.3%) | 1,948 (3.9%) | 2,467 (4.9%) |  |
| Others | 1,831 (3.6%) | 1,836 (3.6%) | 1,933 (3.8%) | 2,282 (4.5%) | 3,530 (7.0%) |  |
| Missing (n) | 232 | 181 | 198 | 223 | 314 |  |
| **TDI** | -1.2 (3.2) | -1.4 (3.0) | -1.5 (3.0) | -1.5 (3.0) | -1.2 (3.2) | <0.001<sup>a</sup> |
| Missing (n) | 55 | 59 | 63 | 60 | 74 |  |
| **BMI** (kg/m2) | 29.8 (4.8) | 28.4 (4.8) | 27.3 (4.6) | 26.4 (4.3) | 25.4 (4.1) | <0.001<sup>a</sup> |
| Missing (n) | 225 | 192 | 172 | 153 | 207 |  |
| **Smoking status** (n%) |  |  |  |  |  | <0.001<sup>b</sup> |
| Never | 22,714 (45.1%) | 26,033 (51.7%) | 27,887 (55.3%) | 29,650 (58.8%) | 32,321 (64.2%) |  |
| Previous | 20,225 (40.2%) | 18,554 (36.8%) | 17,442 (34.6%) | 16,228 (32.2%) | 14,274 (28.3%) |  |
| Current | 7,392 (14.7%) | 5,806 (11.5%) | 5,080 (10.1%) | 4,537 (9.0%) | 3,778 (7.5%) |  |
| Missing (n) | 297 | 235 | 219 | 212 | 254 |  |
| **Alcohol status** (n%) |  |  |  |  |  | <0.001<sup>b</sup> |
| Never | 1,816 (3.6%) | 1,893 (3.7%) | 1,857 (3.7%) | 2,169 (4.3%) | 3,234 (6.4%) |  |
| Previous | 1,896 (3.8%) | 1,665 (3.3%) | 1,615 (3.2%) | 1,636 (3.2%) | 2,085 (4.1%) |  |
| Current | 46,786 (92.6%) | 46,957 (93.0%) | 47,063 (93.1%) | 46,722 (92.5%) | 45,139 (89.5%) |  |
| Missing (n) | 130 | 113 | 93 | 100 | 169 |  |
| **Physical activity** (n%) |  |  |  |  |  | <0.001<sup>b</sup> |
| Low | 9,713 (23.9%) | 8,146 (20.1%) | 7,398 (18.2%) | 6,850 (16.7%) | 6,576 (15.8%) |  |
| Moderate | 16,280 (40.1%) | 16,700 (41.1%) | 16,484 (40.5%) | 16,564 (40.4%) | 16,506 (39.7%) |  |
| High | 14,630 (36.0%) | 15,739 (38.8%) | 16,798 (41.3%) | 17,614 (42.9%) | 18,522 (44.5%) |  |
| Missing (n) | 10,005 | 10,043 | 9,948 | 9,599 | 9,023 |  |

Abbreviations: omega-6%, omega-6 fatty acids to total fatty acids percentage; TDI, Townsend deprivation index; BMI, body mass index.
<sup>a</sup> All variables measured at baseline are presented as mean (SD) unless otherwise specified.<sup>b</sup> From the ANOVA test for continuous variables.<sup>c</sup> From the Pearson's Chi-squared test for categorical variables.

### Associations of plasma omega-6, omega-3, and their ratio with cancer risk

The findings for the associations of plasma omega-6% and omega-3% with the incidence of overall and site-specific cancer are shown in Figure 1, with more detailed information in Tables 2 and 3. In the main models with continuous omega-6% and omega-3%, each standard deviation (SD) increase in the percentage was associated with a 2% (HR per SD = 0.98, 95% CI = 0.96-0.99, p < 0.01) and 1% (HR per SD = 0.99, 95% CI = 0.97 – 1.00, p = 0.03) decline in risk of overall cancer for omega-6% and omega-3%, respectively. Additionally, categorizing omega-6% and omega-3% into quintiles revealed that higher concentrations were linked to a decreased overall cancer risk, with a significant trend observed for both omega-6% and omega-3% (p_trend_ < 0.05).

**Figure 1.**
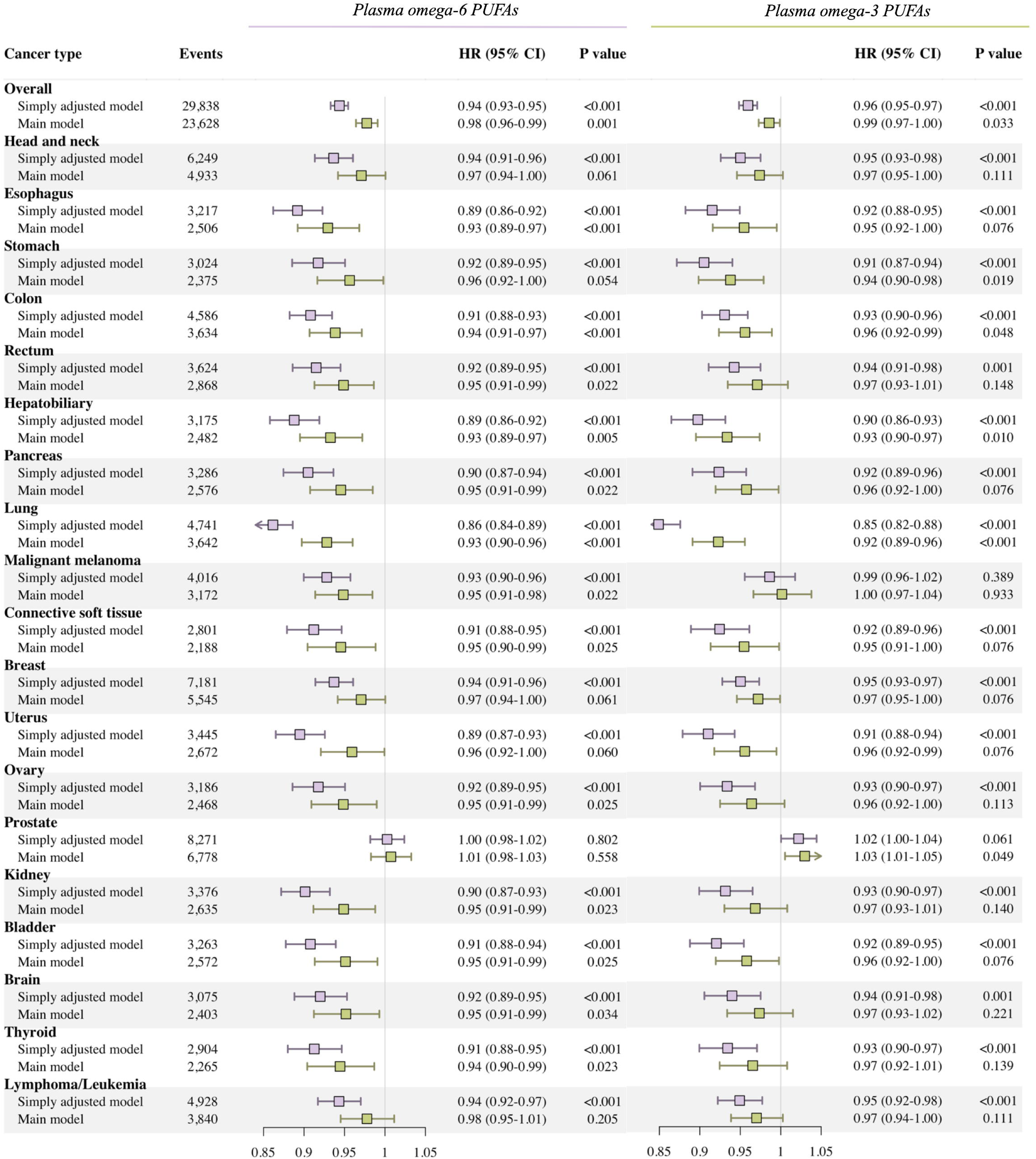
Risk estimates of the incidence of overall cancer and 19 cancer sites for 1-SD increase of plasma omega-6% and omega-3%, for simply adjusted and main models. The results from simply adjusted models revealed the associations stratified by age and sex in the general cohort. The main models were adjusted for general covariates, including ethnicity (classified into White, Black, Asian, Others), Townsend deprivation index (continuous), assessment Center, BMI (kg/m2; continuous), smoking status (categorized as never, previous, current), alcohol intake status (categorized as never, previous, current), and physical activity (classified as low, moderate, high). P values were corrected for the multiple testing of 19 site-specific cancers.

**Table 2.**
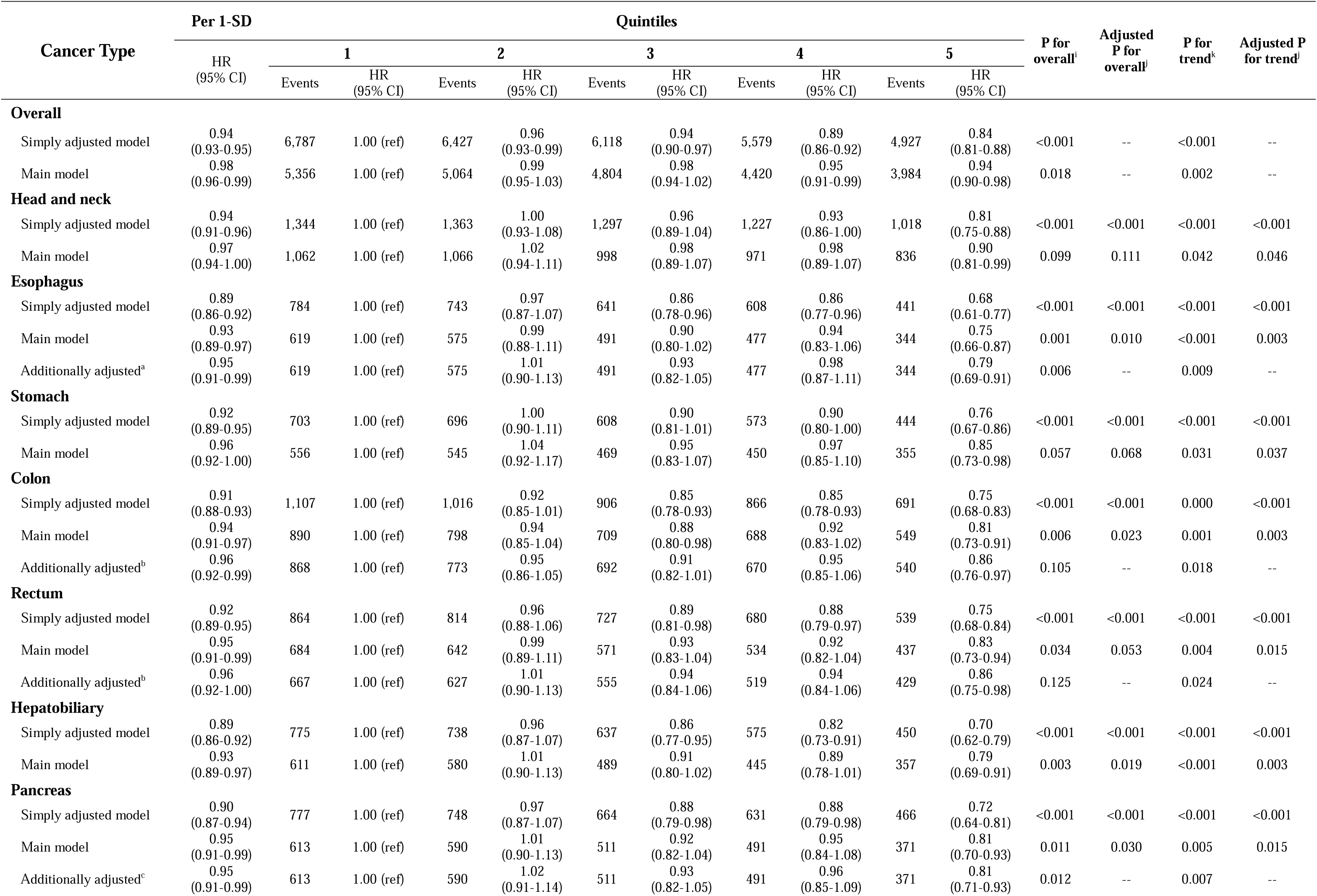
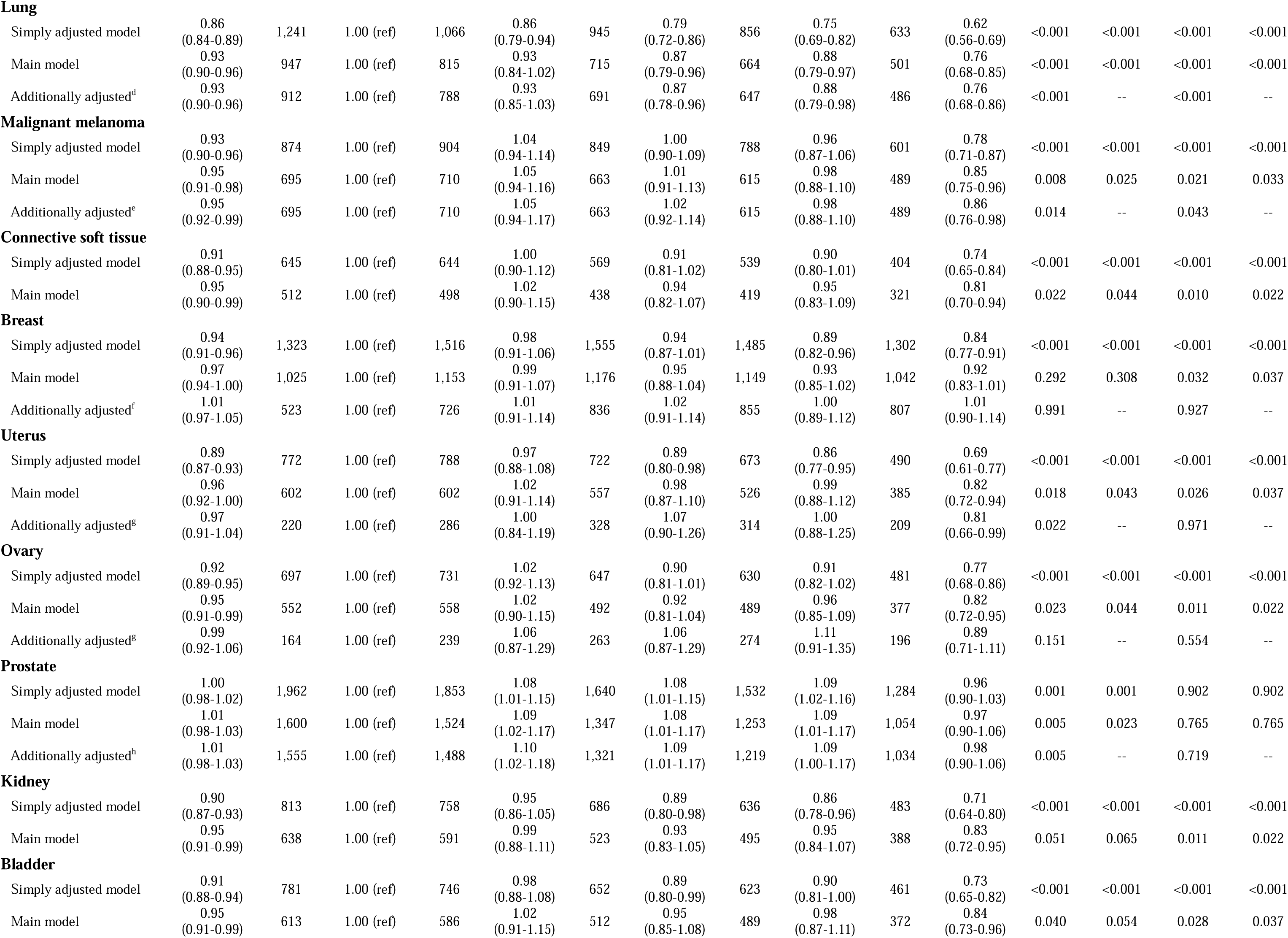
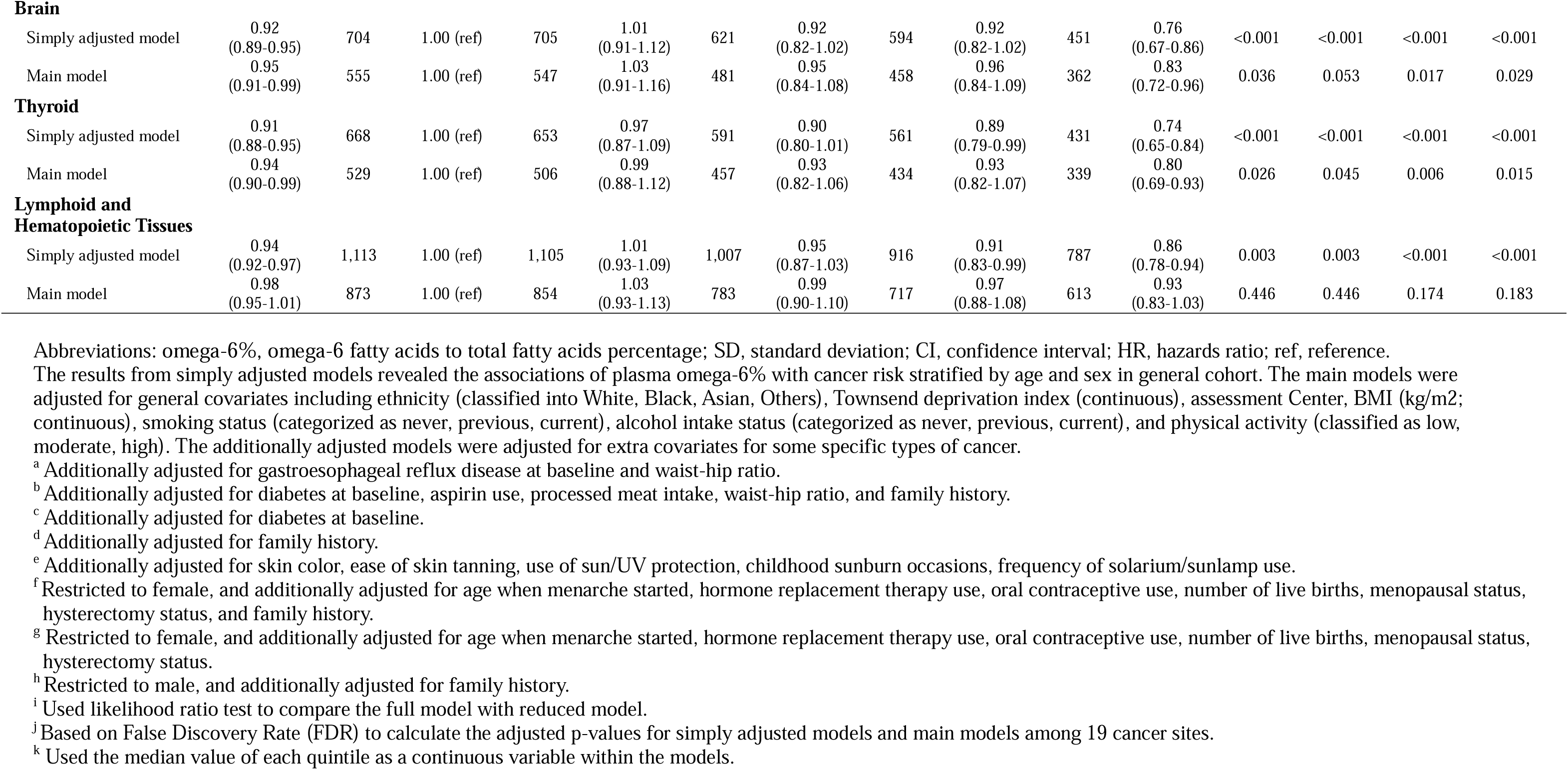
Associations of the plasma omega-6% with the incidence of overall cancer and 19 cancer sites in the UK Biobank.

**Table 3.**
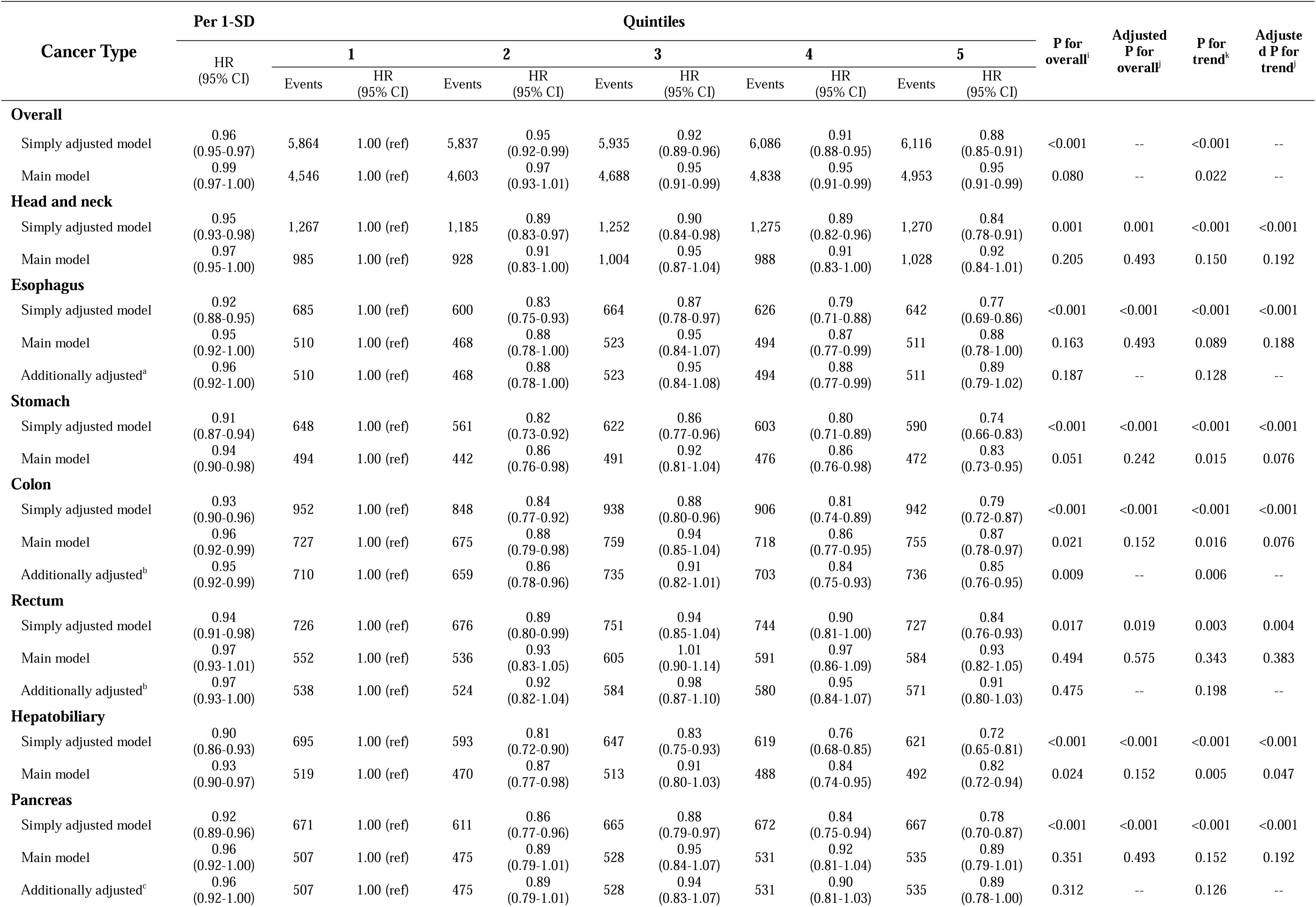
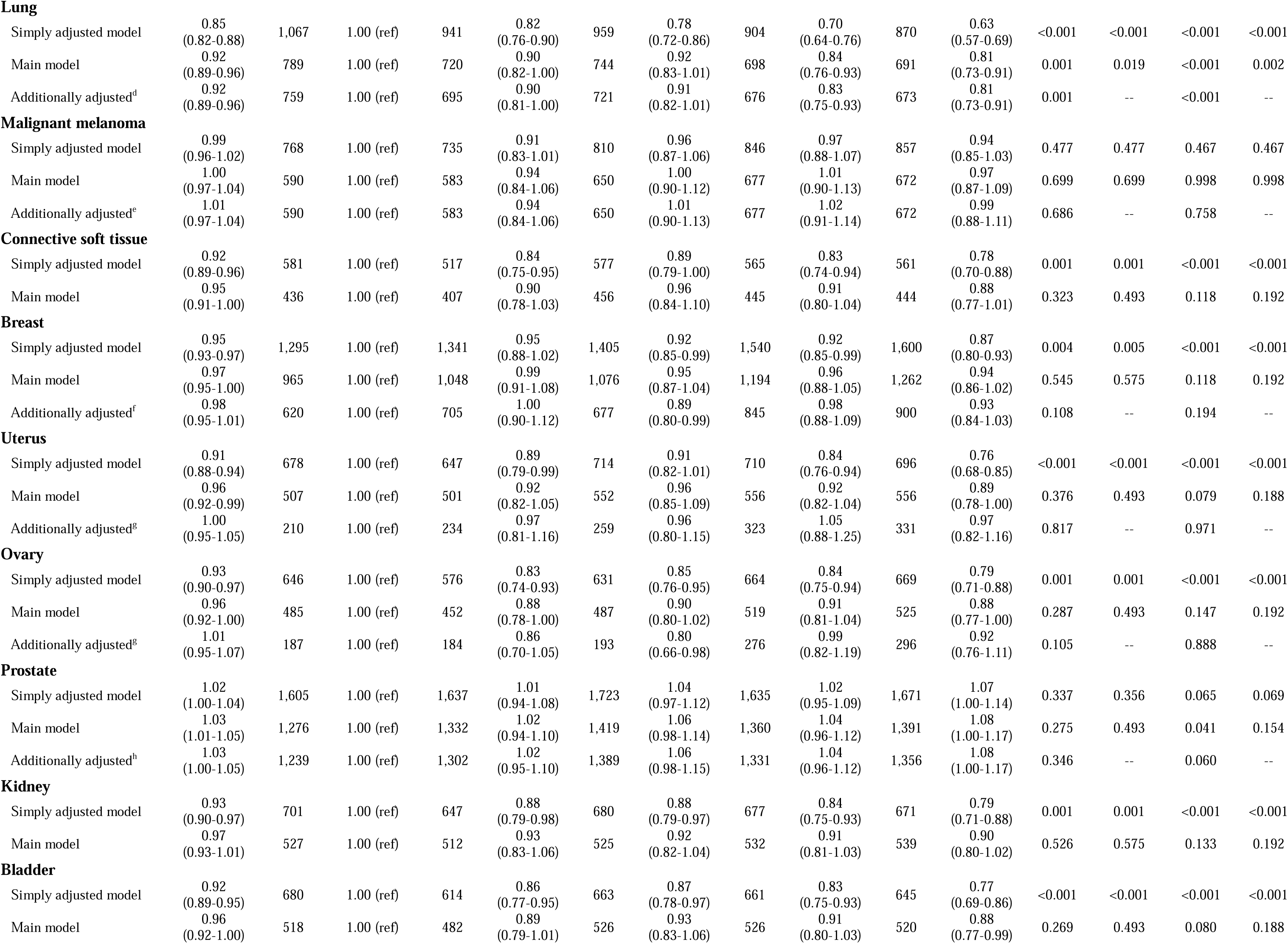
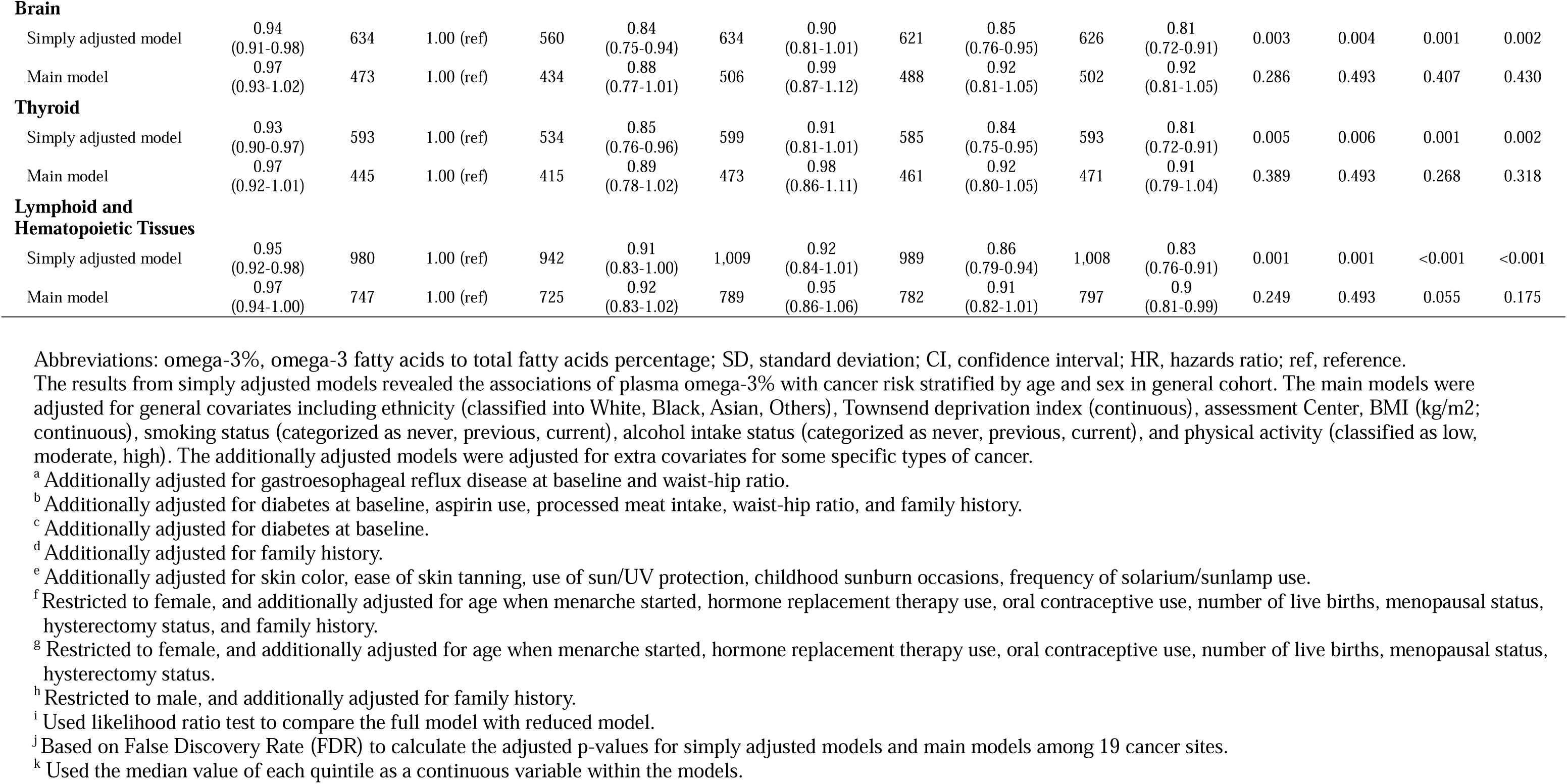
Associations of the plasma omega-3% with the incidence of overall cancer and 19 cancer sites in the UK Biobank.

We performed similar analyses for 19 site-specific cancers. In the main models with continuous exposure, omega-6% was inversely associated with the risk of 13 site-specific cancers (corrected p < 0.05, Figure 1). If considering the trend across the quintiles in the main models, all but two site-specific cancers had inverse associations with omega-6%. The two exceptions were prostate cancer and malignant neoplasms of lymphoid and hematopoietic tissues (corrected p_trend_ < 0.05, Table 2). As for omega-3%, only five site-specific cancers had significant associations in the main analysis with continuous exposure (Figure 1), and the trend analysis across quintiles did not reveal additional significant associations (Table 3). Cancers at four sites, including stomach, colon, hepatobiliary tract, and lung, were inversely associated with both omega-6% and omega-3%. Only one site-specific cancer, prostate cancer, was associated with omega-3% (HR per SD = 1.03, 95% CI = 1.01 – 1.05, corrected p = 0.049) but not omega-6% (HR per SD = 1.01, 95% CI = 0.98 – 1.03, corrected p = 0.56). In the sensitivity analysis of 10 site-specific cancers by additionally adjusting for site-specific covariates, most of the above-mentioned significant associations remained, except the associations of omega-6% with cancers at breast, uterus, and ovary (Figure 2). In summary, we counted associations that were statistically significant in the main models with either continuous exposure analysis or trend analysis, and that remain significant after adjusting for additional site-specific covariates when appropriate. There were 14 site-specific cancers associated with omega-6% and five with omega-3%, with an overlap of four between these two groups. Only four site-specific cancers (i.e., ovary, breast, uterus, and lymphoid and hematopoietic tissues) were not associated with either omega-3% or omega-6%.

**Figure 2.**
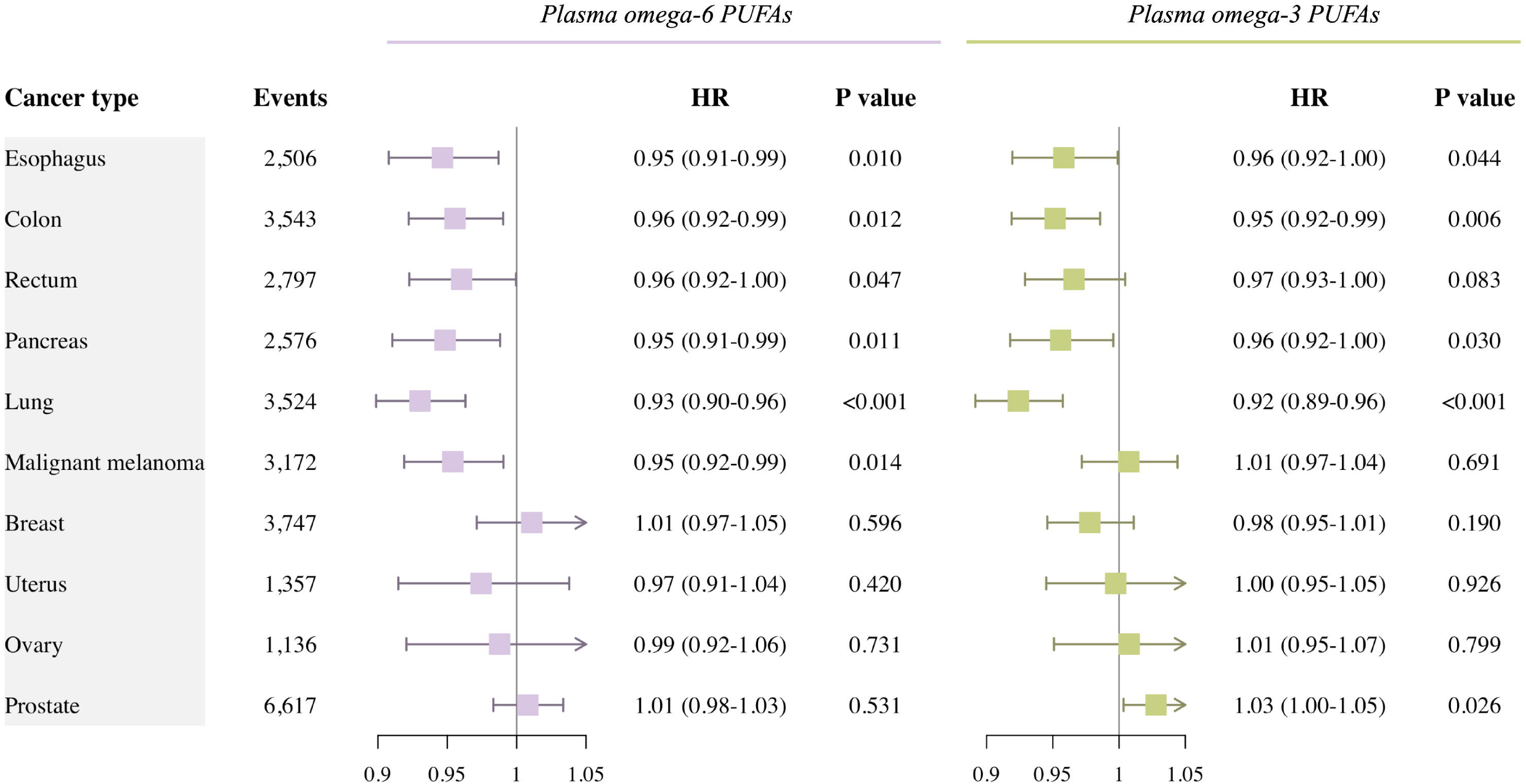
Risk estimates of the incidence of overall cancer and specific cancer sites for 1-SD increase of plasma omega-6% and omega-3%, for additionally adjusted models. For esophagus cancer, additionally adjusted for gastroesophageal reflux disease at baseline and waist-hip ratio. For colon cancer and rectum cancer, additionally adjusted for diabetes at baseline, aspirin use, processed meat intake, waist-hip ratio, and family history. For pancreas cancer, additionally adjusted for diabetes at baseline. For lung cancer, additionally adjusted for family history. For malignant melanoma cancer, additionally adjusted for skin color, ease of skin tanning, use of sun/UV protection, childhood sunburn occasions, frequency of solarium/sunlamp use. For breast cancer, restricted to female, and additionally adjusted for age when menarche started, hormone replacement therapy use, oral contraceptive use, number of live births, menopausal status, hysterectomy status, and family history. For uterus and ovary cancer, restricted to female, and additionally adjusted for age when menarche started, hormone replacement therapy use, oral contraceptive use, number of live births, menopausal status, hysterectomy status. For prostate cancer, restricted to male, and additionally adjusted for family history.

We also conducted analysis of the omega-6/omega-3 ratio (Table S3, Figure S1). A higher omega-6/omega-3 ratio was associated with a higher overall cancer risk (p_trend_ = 0.038). A total of three site-specific cancers showed evidence of positive associations with the ratio. Every SD increment in the ratio was associated with a 2% increase in the risk of rectum cancer, and the association remained unchanged after additionally controlling for site-specific covariates (per SD HR = 1.02, 95% CI = 1.01 – 1.03, p = 0.003). When examining trends across quintiles, lung cancer was significant in the main model (corrected p_trend_ = 0.011) and remained significant after adjusting for additional covariates (p_trend_ < 0.001). Colon cancer was significant in the additionally adjusted model (p_trend_ = 0.015).

In addition to the trend analysis across quintiles, we assessed if there were any differences across the association effect sizes of quintiles by applying likelihood ratio tests. Most of the PUFAs-cancer relationships with significant trends were also statistically significant in the overall likelihood ratio tests. On the other hand, there were two pairs of relationships whose trend analyses were not significant, but their overall tests were. The most notable pair was omega-6% and prostate cancer (additionally adjusted model, p_trend_ = 0.72, p_overall_ = 0.005). The association estimates across the quintiles support the presence of a nonlinear relationship: Quintile 2 (HR = 1.10, 95% CI = 1.02 – 1.18), Quintile 3 (HR = 1.09, 95% CI = 1.01 – 1.17), Quintile 4 (HR = 1.09, 95% = 1.00 – 1.17), and Quintile 5 (HR = 0.98, 95% CI = 0.90-1.06). The other pair was omega-6% and uterus cancer (additionally adjusted model, p_trend_ = 0.97, p_overall_ = 0.022), and there was an inverse association in the Quintile 5 (HR = 0.81, 95% CI = 0.66-0.99).

### Stratified analyses for plasma omega-6 and omega-3 fatty acids

Stratified analyses were conducted to assess potential effect modifications by age, sex, TDI, BMI, smoking status, alcohol consumption status, and physical activity, as shown in Table 4. The observed inverse associations of plasma omega-6% with overall cancer risk appeared to be notably more pronounced in the younger age group (p for interaction <0.001) and in females (p for interaction = 0.006), with no apparent modification by the remaining potential stratification variables. Moreover, the estimated inverse associations of plasma omega-3% with overall cancer risk demonstrated a tendency to be stronger in the older group (p for interaction < 0.001), in males (p for interaction = 0.002), and in current smokers (p for interaction = 0.017).

**Table 4.**
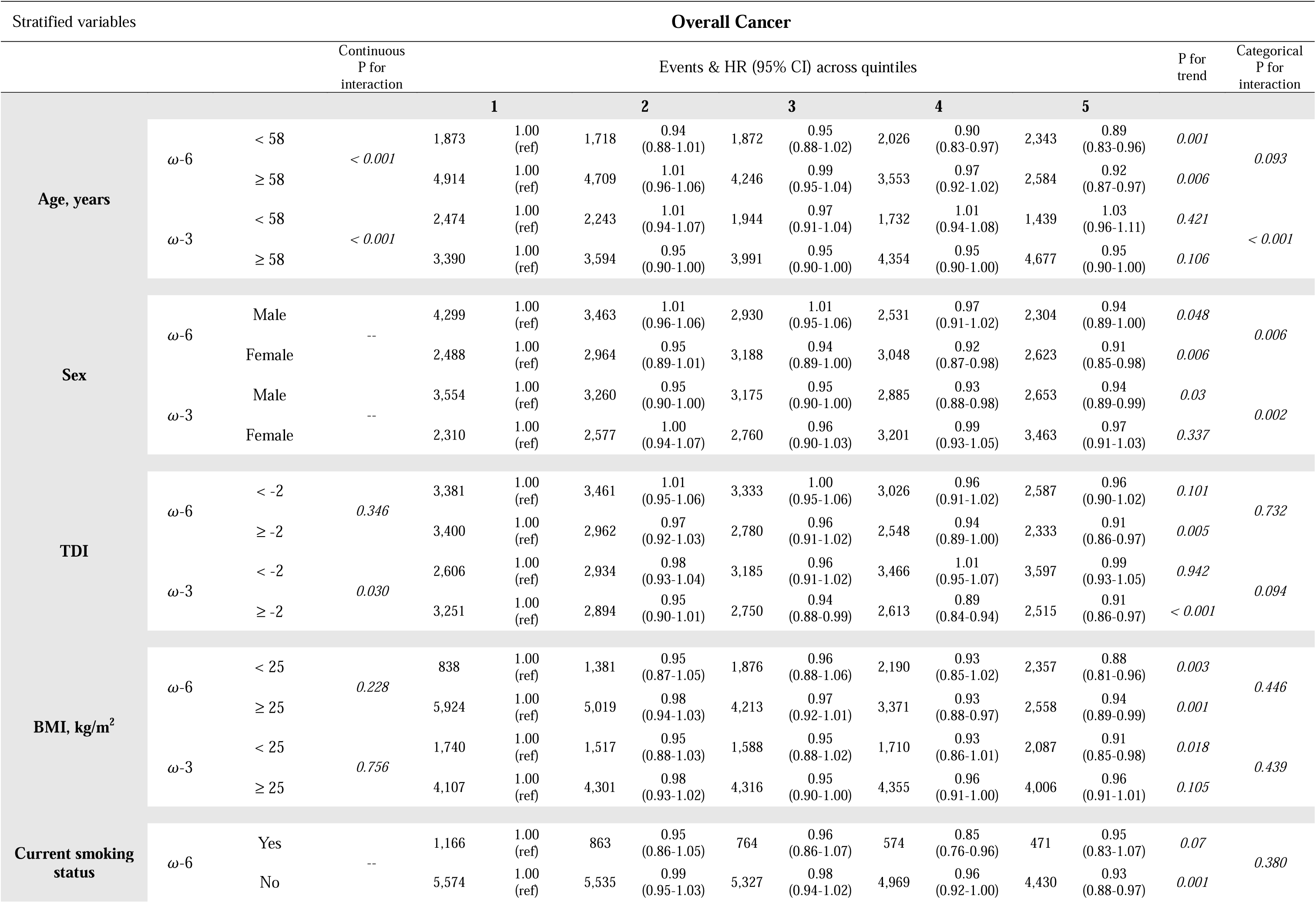
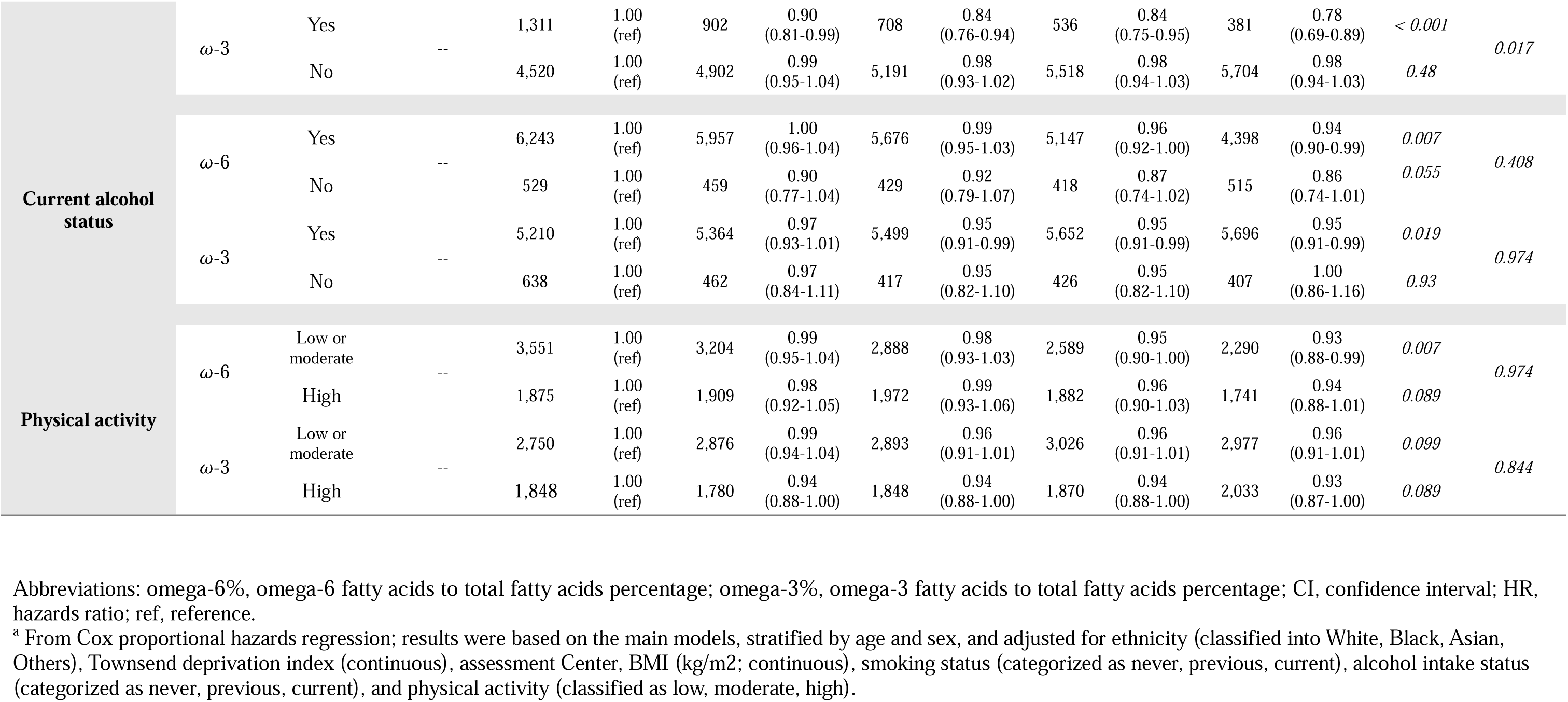
Risk estimates^a^ of plasma omega-6% and omega-3% with incidence of overall cancer, stratified by potential risk factors, in the UK Biobank Study (n = 253,138)

### Restricted cubic spline analysis

In the restricted cubic spline analysis, it is noteworthy that significant inverse associations were observed for omega-6% and omega-3% with the overall cancer incidence (p < 0.05 for both variables, as shown in Figure S2). Moreover, potential nonlinearity was identified for the relationship between omega-3% and overall cancer incidence (p < 0.05). This finding suggests that the protective effect of omega-3 PUFAs may exhibit enhanced efficacy at the lower concentration level. Due to the possible presence of a nonlinear association between omega-6% and prostate cancer, we further performed cubic spline analysis for prostate cancer. We found evidence of nonlinearity between omega-6% and prostate cancer, with the intermediate level of omega-6% associated with the highest risk (p = 0.02, Figure S3).

### Sensitivity analyses

In order to evaluate whether the associations between plasma omega-6% and overall cancer risk might undergo modification by omega-3%, or vice versa, both omega-6% and omega-3% were simultaneously integrated into the same models (as detailed in Table S4 and Table S5). The correlation between plasma omega-6% and omega-3% was relatively low, with r = –0.12 (p < 0.01). After their inclusion in the same models, the associations of both plasma omega-6% and omega-3% with overall cancer risk remained statistically significant. The results for DHA% and LA% were consistent with those for omega-3% and omega-6%, respectively (as detailed in Table S6 and Table S7). Additionally, when we excluded participants who experienced cancer or death within the first year or the first three years of follow-up, the outcomes remained unchanged (as detailed in Table S8 and Table S9). It is worth noting that the baseline characteristics were comparable between participants with and without exposure information, as evidenced by Table S10.

## Discussion

Our population-based prospective cohort study in UK Biobank revealed that higher plasma omega-6% and omega-3% were both associated with a lower incidence of overall cancer. The overall association effect sizes in the main model were 2% and 1% reductions per SD of omega-6% and omega-3%, respectively. The association of omega-6% with cancer risk was independent of most risk factors examined, including TDI, BMI, smoking status, alcohol status, and physical activity. The observed inverse associations of plasma omega-6% appeared to be notably more pronounced in the younger age group and in women. On the other hand, the inverse associations of plasma omega-3% with overall cancer incidence were stronger in the older age group, in men, and in current smokers. The inverse associations of omega-6% and omega-3% with overall cancer incidence were robust to a list of sensitivity analyses. In terms of the incidence of 19 site-specific cancers, 14 were associated with omega-6% and five with omega-3%, all exhibiting inverse associations (3% – 7% reduced risk per SD of omega-6%; 5% – 8% reduced risk per SD of omega-3%), with the exception that prostate cancer was positively associated with omega-3% (3% increased risk). Only four site-specific cancers (i.e., ovary, breast, uterus, and lymphoid and hematopoietic tissues) were not associated with either omega-3% or omega-6%.

Despite a large number of studies, the links between PUFAs, especially omega-6 PUFAs, and the incidence of overall cancer remain ill-defined. Most existing studies examined dietary PUFAs or supplements, instead of circulating biomarkers. A 2019 prospective cohort study found no significant associations of omega-3 or omega-6 PUFA intakes with the overall cancer incidence [19]. A 2020 meta-analysis of randomized trials showed that increasing dietary long-chain omega-3 PUFAs had little or no effects on overall cancer diagnosis or cancer death, while the effects of increasing dietary omega-6 PUFAs were unclear because the evidence was of very low quality [2]. A 2022 meta-analysis of observational studies revealed that fish intake and marine omega-3 PUFA intake were associated with lower mortality in patients with overall cancer [7]. Of note, a 2020 meta-analysis of prospective studies showed that the blood level of omega-6 PUFAs (highest vs. lowest category RR = 0.92, 95% CI = 0.86 – 0.98), but not their intake, was inversely associated with overall cancer risk [10]. They also found that the protective association was stronger in women than in men, consistent with our findings. In the context of UK Biobank, a 2021 prospective study demonstrated that regular fish oil supplementation was associated with a lower incidence of overall cancer, but only in participants who consumed fatty fish less than two times per week (HR = 0.96, 95% CI = 0.94 – 0.99), not in those who consumed more than twice per week (HR = 1.01, 95% CI = 0.95 – 1.07). Their subgroup analysis further unraveled that men were more likely to gain benefits from fish oil supplementation than women [15]. Consistently, our study found that the plasma level of omega-3 PUFAs was inversely associated with overall cancer incidence and that the association was only significant in men. Moreover, a 2023 study of circulating PUFAs and cancer mortality by our group revealed that both plasma omega-3 and omega-6 PUFAs were inversely associated with cancer mortality (highest vs. lowest quintile HR = 0.75, 95% = 0.65 – 0.87; HR = 0.80, 95% CI = 0.68 – 0.92; respectively) [17]. Overall, our findings provide support for possible small net protective roles of omega-3 and omega-6 PUFAs in the development of new cancer incidence. Our study also suggests that the usage of circulating blood biomarkers captures different aspects of dietary intake, reduces measurement errors, and thus enhances statistical power. The differential effects of omega-6% and omega-3% in age and sex subgroups warrant future investigation.

In our study, we observed site-specific associations of omega-3 PUFAs with cancer incidence. A higher plasma level of omega-3 PUFAs was associated with a significant reduction in the incidence of digestive system cancers (including colon, stomach, and hepatobiliary tract) and lung cancer. However, it appeared to be linked to an increased risk of prostate cancer. The observed protective associations between plasma omega-3 PUFAs and the incidence of digestive system cancers and lung cancer are consistent with recent studies of fish oil supplementation and dietary intake in UK Biobank [13–15]. Regular fish oil supplementation was associated with lower incidence of colon cancer (HR = 0.88, 95% CI = 0.8-0.98), hepatobiliary cancer (HR = 0.72, 95% CI = 0.58-0.91), and lung cancer (HR = 0.87, 95% CI = 0.78-0.96) [15]. Another independent analysis of UK Biobank data revealed a 44% lower risk of liver cancer incidence among fish oil users [14]. Dietary intake of omega-3 PUFAs was associated with an 18% decreased risk in lung cancer incidence (HR=0.82, 95% CI= 0.73-0.93; per 1g/d) [13]. Notably, some studies and meta-analyses did not find significant associations of dietary omega-3 PUFAs and fish oil supplementation with colorectal cancer [3, 20, 21]. However, a recent meta-analysis showed that while the dietary intake of omega-3 PUFAs was not associated with the colorectal cancer risk (relative risk, RR = 0.97, 95% CI = 0.90 – 1.04 for the highest versus lowest category), the blood level of omega-3 PUFAs was associated with a lower risk (RR = 0.79, 95% CI = 0.64 – 0.98) [11]. Regarding prostate cancer, most studies did not find significant associations with dietary intake or blood level of omega-3 PUFAs [3, 15, 22–24]. However, the few statistically significant findings suggest that dietary intake of alpha-linolenic acid (ALA) was associated with a lower prostate cancer risk, while both dietary intake and blood level of DHA were associated with a higher risk [3, 23, 25]. Our study found that plasma omega-3% and DHA% were both positively associated with the risk of prostate cancer. Further studies are warranted to explore the roles of individual omega-3 PUFAs in the etiology of prostate cancer.

In our investigation of omega-6 PUFAs, we observed inverse associations of plasma omega-6 PUFAs with 14 site-specific cancers at head and neck, esophagus, stomach, colon, rectum, hepatobiliary tract, pancreas, lung, malignant melanoma, connective soft tissue, kidney, bladder, brain, and thyroid. Moreover, an increased omega-6/omega-3 PUFAs ratio was associated with elevated risks of rectum, colon, and lung cancer. Notably, the evidence on the associations between omega-6 PUFAs, the omega-6/omega-3 ratio, and site-specific cancers was limited and exhibited varying results. Two prospective cohort studies did not establish significant links between dietary omega-6 PUFAs and colorectal cancer [20, 21]. However, in agreement with our findings, another prospective cohort study observed that omega-6 PUFA intake was inversely associated with the risk of digestive cancer (including esophagus, liver, stomach, pancreas, and colorectal) (highest vs. lowest quintile HR = 0.56, 95% CI = 0.32 – 0.97) or colorectal cancer alone (HR = 0.43, 95% CI = 0.22 – 0.83)(19). Also consistent with our results, a prospective cohort study based on UK Biobank indicated a modest protective effect of dietary omega-6 PUFAs against lung cancer (HR = 0.98, 95% CI = 0.96-0.99; per 1g/d) [13]. A systematic review and meta-analysis of eight previous studies also found no apparent association between dietary omega-6 PUFAs and prostate cancer [23], in line with our findings from trend analysis. However, we did find evidence for the possible presence of a nonlinear relationship, with intermediate levels of omega-6% associated with the highest risk of prostate cancer. There were three site-specific cancers at breast, uterus, and ovary that were inversely associated with plasma omega-6% in our main models, but these associations disappeared after controlling for site-specific covariates, such as age of menarche, hormone replacement therapy use, oral contraceptive use, number of live births, menopausal status, and hysterectomy status. A previous meta-analysis of prospective studies did observe an inverse association of the blood omega-6 level with breast cancer (highest vs. lowest category RR = 0.87; 95% CI: 0.77–0.98) [10]. Our study indicated that the consideration of site-specific covariates is critical in interpreting associations.

This study has several strengths. The major strength was the prospective population-based study design in UK Biobank, which provides a large sample size, long duration of follow-up, and detailed information on potential confounding variables. We used the objective measurements of PUFA biomarkers in plasma instead of the estimated dietary intakes from self-reported questionnaires, which increases the accuracy of exposure assessment. Moreover, the cancer incidence data were acquired through cancer registries to reduce selection bias. This approach ensures a more representative sample, as these registries comprehensively cover a wide range of demographics and cancer types, and adhere to standardized data collection protocols, thereby enhancing the reliability and generalizability of our findings [26]. Furthermore, we adopted the FDR approach when investigating site-specific cancers, to address the issue of increasing false positives from multiple comparisons. In several sensitivity analyses, most of the associations remain materially unchanged, indicating the robustness of our results.

Some potential limitations warrant consideration in the interpretation of our findings. First, despite previous indications of the representativeness of UK Biobank in sociodemographic and health-related characteristics of the UK population, the potential for selective bias persists [27, 28]. Notably, the participant sample skewed heavily toward European ancestry and White ethnicity, necessitating caution in generalizing results across diverse ancestral backgrounds and ethnicities. Secondly, while we adjusted for multiple potential confounding variables in our model, the inherent limitations of observational studies preclude the complete elimination of inaccuracies in measurements, unmeasured variables, and interdependencies among factors. Thirdly, the number of events was small for some specific cancer sites, which may lead to the limited statistical power of our study. Fourthly, our study focused on total omega-3 and omega-6 PUFAs. There are only two individual PUFAs measured in the UK Biobank cohort, LA and DHA. We showed that the associations of LA% mirrored those of omega-6%, while DHA% mirrored omega-3%. Future studies into other individual PUFAs are needed. Lastly, despite the relative homogeneity of the sample, individual genetics have not been taken into account. Future studies are warranted to examine if specific genetic variants or composite genetic scores modify the associations of circulating PUFAs with overall or site-specific cancers.

## Conclusion

In our UK Biobank prospective cohort study, elevated levels of plasma omega-6 and omega-3 PUFAs were linked to reduced overall cancer risk, while a higher omega-6/omega-3 ratio was associated with increased risk. The associations of omega-6 PUFAs were stronger in the younger age group and in women, while the associations of omega-3 PUFAs were more prominent in the older group, in men, and in current smokers. Our findings extended to the inverse associations of plasma omega-6 and omega-3 PUFAs with 14 site-specific cancers. One notable exception to this trend of protective association was between omega-3 PUFAs and prostate cancer. Our study laid a solid foundation for future mechanistic studies into the roles of PUFAs in the etiology of various cancers. It also provided insights into the development of cancer prevention strategies by managing circulating PUFAs.

## Additional Information

## Acknowledgements

This research has been conducted using the UK Biobank Resource under Application Number 48818. This work uses data provided by patients and collected by the NHS as part of their care and support. These data are copyrighted, 2022, NHS England. Reused with the permission of the NHS England and UK Biobank. All rights reserved. This research used data assets made available by National Safe Haven as part of the Data and Connectivity National Core Study, led by Health Data Research UK in partnership with the Office for National Statistics and funded by UK Research and Innovation (grant ref MC_PC_20058). We would like to express our heartfelt gratitude to the UK Biobank participants and administrative staff.

## Authors’ contributions

YZ performed data analysis, prepared visualizations, and wrote the original draft of the manuscript. YS and SS contributed to the data analysis. NKK and JTB contributed to the interpretations of results. YS and KY contributed equally to this project and should be considered co-corresponding authors. They jointly designed and supervised the project. All authors critically edited the manuscript for important intellectual content. The corresponding author (KY) attests that all listed authors meet the authorship criteria and that no others meeting the criteria were omitted.

## Ethics approval and consent to participate

The UK Biobank received ethical approval from the research ethics committee (reference ID: 11/ NW/0382). Written informed consent was obtained from participants.

## Consent for publication

Not applicable.

## Data availability

The datasets analyzed during the current study are available from the UK Biobank through an application process (www.ukbiobank.ac.uk/).

