## Supplementary Materials for "Associations of plasma omega-6 and omega-3 fatty acids with overall and 19 site-specific cancers: a population-based cohort study in UK Biobank"

Table S1. Extra covariates to specific subtypes of cancer in additionally adjusted models.

| **Cancer subtype** | **Additionally adjusted covariates** |
| --- | --- |
| Esophagus | gastroesophageal reflux disease at baseline and waist-hip ratio |
| Colon and rectum | diabetes at baseline, aspirin use, processed meat intake, waist-hip ratio, and family history |
| Pancreas | diabetes at baseline |
| Lung | family history |
| Malignant melanoma | skin color, ease of skin tanning, use of sun/UV protection, childhood sunburn occasions, frequency of solarium/sunlamp use |
| Breast | Restricted to female, and additionally adjusted for age when menarche started, hormone replacement therapy use, oral contraceptive use, number of live births, menopausal status, hysterectomy status, and family history |
| Uterus and ovary | Restricted to female, and additionally adjusted for age when menarche started, hormone replacement therapy use, oral contraceptive use, number of live births, menopausal status, hysterectomy status |
| Prostate | Restricted to male, and additionally adjusted for family history |

Table S2. Baseline characteristics of included participants by quintiles of the plasma omega-3% (n = 253,138)

|  | **Omega-3% quintiles** | | | | | |  |
| --- | --- | --- | --- | --- | --- | --- | --- |
| **Characteristics**^a^ | **1** (median = 2.7)  (n = 50,628) | **2** (median = 3.5) (n = 50,628) | **3** (median = 4.2) (n = 50,628) | **4** (median = 4.9)  (n = 50,627) | **5** (median = 6.3) (n = 50,627) | | **p-value** |
| **Age** (years) | 54.2 (8.2) | 55.2 (8.2) | 56.3 (8.1) | 57.4 (7.8) | | 58.8 (7.3) | <0.001^a^ |
| **Gender** (male%) | 55.9 | 51.9 | 48.4 | 42.7 | | 37.0 | <0.001^b^ |
| **Ethnicity**(n%) |  |  |  |  | |  |  |
| White | 46,254 (91.8%) | 46,196 (91.6%) | 46,223 (91.7%) | 46,126 (91.5%) | | 45,423 (90.1%) | <0.001^b^ |
| Black | 269 (0.5%) | 276 (0.5%) | 241 (0.5%) | 264 (0.5%) | | 327 (0.6%) |  |
| Asian | 1,676 (3.3%) | 1,737 (3.4%) | 1,767 (3.5%) | 1,809 (3.6%) | | 1,990 (3.9%) |  |
| Others | 2,166 (4.3%) | 2,201 (4.4%) | 2,170 (4.3%) | 2,209 (4.4%) | | 2,666 (5.3%) |  |
| Missing (n) | 263 | 218 | 227 | 219 | | 221 |  |
| **TDI** | -0.8 (3.2) | -1.2 (3.1) | -1.4 (3.0) | -1.6 (3.0) | | -1.7 (3.0) | <0.001^a^ |
| Missing (n) | 66 | 81 | 56 | 59 | | 49 |  |
| **BMI** (kg/m2) | 27.8 (5.3) | 27.9 (4.9) | 27.7 (4.7) | 27.3 (4.5) | | 26.6 (4.2) | <0.001^a^ |
| Missing (n) | 244 | 173 | 207 | 163 | | 162 |  |
| **Smoking status** (n%) |  |  |  |  | |  | <0.001^b^ |
| Never | 26,466 (52.5%) | 27,174 (53.9%) | 27,563 (54.7%) | 28,163 (55.9%) | | 29,239 (58.0%) |  |
| Previous | 15,202 (30.2%) | 16,842 (33.4%) | 17,852 (35.4%) | 18,377 (36.5%) | | 18,450 (36.6%) |  |
| Current | 8,711 (17.3%) | 6,369 (12.6%) | 4,974 (9.9%) | 3,843 (7.6%) | | 2,696 (5.4%) |  |
| Missing (n) | 249 | 243 | 239 | 244 | | 242 |  |
| **Alcohol status** (n%) |  |  |  |  | |  | <0.001^b^ |
| Never | 2,743 (5.4%) | 2,277 (4.5%) | 2,020 (4.0%) | 1,972 (3.9%) | | 1,957 (3.9%) |  |
| Previous | 2,492 (4.9%) | 1,829 (3.6%) | 1,561 (3.1%) | 1,512 (3.0%) | | 1,503 (3.0%) |  |
| Current | 45,233 (89.6%) | 46,418 (91.9%) | 46,925 (92.9%) | 47,045 (93.1%) | | 47,046 (93.1%) |  |
| Missing (n) | 160 | 104 | 122 | 98 | | 121 |  |
| **Physical activity** (n%) |  |  |  |  | |  | <0.001^b^ |
| Low | 8,155 (20.2%) | 8,109 (19.9%) | 7,888 (19.3%) | 7,691 (18.7%) | | 6,840 (16.6%) |  |
| Moderate | 15,503 (38.4%) | 16,073 (39.4%) | 16,622 (40.6%) | 17,197 (41.8%) | | 17,139 (41.5%) |  |
| High | 16,727 (41.4%) | 16,571 (40.7%) | 16,442 (40.1%) | 16,266 (39.5%) | | 17,297 (41.9%) |  |
| Missing (n) | 10,243 | 9,875 | 9,676 | 9,473 | | 9,351 |  |

Abbreviations: omega-3%, omega-3 fatty acids to total fatty acids percentage; TDI, Townsend deprivation index; BMI, body mass index.

^a^ All variables measured at baseline are presented as mean (SD) unless otherwise specified.

^b^ From the ANOVA test for continuous variables.

^c^ From the Pearson's Chi-squared test for categorical variables.

Table S3. Associations of the plasma omega-6/omega-3 ratio with the incidence of overall cancer and 19 cancer sites in the UK Biobank

| **Cancer Type** | **Per 1-SD** | **Quintiles** | | | | | | | | | | **P for overall**^i^ | **Adjusted P for overall**^j^ | **P for trend**^k^ | **Adjusted P for trend**^j^ |
| --- | --- | --- | --- | --- | --- | --- | --- | --- | --- | --- | --- | --- | --- | --- | --- |
|  | HR  (95% CI) | **1** | | **2** | | **3** | | **4** | | **5** | |  |  |  |  |
|  |  | Events | HR  (95% CI) | Events | HR  (95% CI) | Events | HR  (95% CI) | Events | HR  (95% CI) | Events | HR  (95% CI) |  |  |  |  |
| **Overall** |  |  |  |  |  |  |  |  |  |  |  |  |  |  |  |
| Simply adjusted model | 1.01  (1.00-1.02) | 6,330 | 1.00 (ref) | 6,232 | 1.03  (0.99-1.07) | 5,888 | 1.01  (0.98-1.05) | 5,757 | 1.04  (1.01-1.08) | 5,631 | 1.08  (1.04-1.12) | <0.001 | -- | <0.001 | -- |
| Main model | 1.01  (1.00-1.02) | 5,101 | 1.00 (ref) | 4,958 | 1.01  (0.97-1.05) | 4,635 | 0.98  (0.94-1.02) | 4,526 | 1.01  (0.97-1.05) | 4,408 | 1.04  (1.00-1.09) | 0.060 | -- | 0.038 | -- |
| **Head and neck** |  |  |  |  |  |  |  |  |  |  |  |  |  |  |  |
| Simply adjusted model | 1.01  (0.99-1.03) | 1,310 | 1.00 (ref) | 1,278 | 1.02  (0.94-1.10) | 1,258 | 1.04  (0.96-1.13) | 1,201 | 1.04  (0.96-1.12) | 1,202 | 1.09  (1.01-1.18) | 0.267 | 0.298 | 0.026 | 0.031 |
| Main model | 1.00  (0.98-1.03) | 1,038 | 1.00 (ref) | 1,016 | 1.02  (0.93-1.11) | 996 | 1.02  (0.94-1.12) | 938 | 1.01  (0.92-1.10) | 945 | 1.06  (0.97-1.16) | 0.774 | 0.774 | 0.250 | 0.316 |
| **Esophagus** |  |  |  |  |  |  |  |  |  |  |  |  |  |  |  |
| Simply adjusted model | 1.01  (0.99-1.03) | 673 | 1.00 (ref) | 658 | 1.03  (0.93-1.15) | 647 | 1.07  (0.96-1.19) | 601 | 1.05  (0.94-1.18) | 638 | 1.20  (1.07-1.34) | 0.025 | 0.090 | 0.002 | 0.006 |
| Main model | 1.01  (0.97-1.04) | 534 | 1.00 (ref) | 525 | 1.03  (0.91-1.16) | 500 | 1.02  (0.90-1.15) | 466 | 1.01  (0.89-1.14) | 481 | 1.10  (0.97-1.25) | 0.567 | 0.769 | 0.155 | 0.265 |
| Additionally adjusted^a^ | 1.01  (0.98-1.04) | 534 | 1.00 (ref) | 525 | 1.02  (0.91-1.16) | 500 | 1.02  (0.90-1.15) | 466 | 1.01  (0.89-1.15) | 481 | 1.11  (0.98-1.26) | 0.542 | -- | 0.127 | -- |
| **Stomach** |  |  |  |  |  |  |  |  |  |  |  |  |  |  |  |
| Simply adjusted model | 1.01  (0.99-1.03) | 625 | 1.00 (ref) | 609 | 1.03  (0.92-1.16) | 612 | 1.09  (0.98-1.22) | 566 | 1.07  (0.96-1.21) | 612 | 1.25  (1.11-1.4) | 0.003 | 0.019 | <0.001 | 0.001 |
| Main model | 1.01  (0.98-1.04) | 499 | 1.00 (ref) | 485 | 1.02  (0.90-1.16) | 474 | 1.03  (0.91-1.17) | 441 | 1.03  (0.90-1.17) | 476 | 1.17  (1.03-1.33) | 0.125 | 0.594 | 0.015 | 0.095 |
| **Colon** |  |  |  |  |  |  |  |  |  |  |  |  |  |  |  |
| Simply adjusted model | 1.01  (0.99-1.03) | 991 | 1.00 (ref) | 937 | 1.01  (0.92-1.10) | 916 | 1.04  (0.95-1.14) | 860 | 1.04  (0.95-1.14) | 882 | 1.15  (1.04-1.26) | 0.038 | 0.090 | 0.002 | 0.006 |
| Main model | 1.01  (0.98-1.04) | 790 | 1.00 (ref) | 766 | 1.02  (0.93-1.13) | 718 | 1.00  (0.91-1.11) | 675 | 1.01  (0.91-1.12) | 685 | 1.10  (0.99-1.23) | 0.349 | 0.636 | 0.079 | 0.265 |
| Additionally adjusted^b^ | 1.01  (0.98-1.03) | 767 | 1.00 (ref) | 749 | 1.03  (0.94-1.14) | 700 | 1.02  (0.92-1.13) | 659 | 1.03  (0.93-1.15) | 668 | 1.15  (1.03-1.28) | 0.123 | -- | 0.015 | -- |
| **Rectum** |  |  |  |  |  |  |  |  |  |  |  |  |  |  |  |
| Simply adjusted model | 1.02  (1.01-1.03) | 776 | 1.00 (ref) | 764 | 1.04  (0.94-1.14) | 719 | 1.02  (0.92-1.13) | 672 | 1.01  (0.91-1.12) | 693 | 1.10  (1.00-1.23) | 0.356 | 0.376 | 0.099 | 0.111 |
| Main model | 1.02  (1.01-1.03) | 622 | 1.00 (ref) | 616 | 1.03  (0.93-1.16) | 566 | 0.98  (0.88-1.10) | 529 | 0.98  (0.87-1.10) | 535 | 1.04  (0.93-1.18) | 0.735 | 0.774 | 0.692 | 0.692 |
| Additionally adjusted^b^ | 1.02  (1.01-1.03) | 610 | 1.00 (ref) | 598 | 1.03  (0.92-1.15) | 549 | 0.98  (0.88-1.10) | 519 | 1.00  (0.88-1.12) | 521 | 1.07  (0.95-1.21) | 0.633 | -- | 0.334 | -- |
| **Hepatobiliary** |  |  |  |  |  |  |  |  |  |  |  |  |  |  |  |
| Simply adjusted model | 1.02  (1.00-1.03) | 660 | 1.00 (ref) | 638 | 1.03  (0.92-1.14) | 639 | 1.08  (0.97-1.21) | 586 | 1.06  (0.94-1.18) | 652 | 1.26  (1.13-1.41) | <0.001 | <0.001 | <0.001 | <0.001 |
| Main model | 1.01  (0.99-1.04) | 524 | 1.00 (ref) | 512 | 1.02  (0.91-1.16) | 490 | 1.02  (0.90-1.16) | 458 | 1.02  (0.90-1.16) | 498 | 1.18  (1.04-1.34) | 0.074 | 0.594 | 0.011 | 0.095 |
| **Pancreas** |  |  |  |  |  |  |  |  |  |  |  |  |  |  |  |
| Simply adjusted model | 1.01  (0.99-1.03) | 708 | 1.00 (ref) | 675 | 1.02  (0.92-1.13) | 665 | 1.06  (0.95-1.18) | 601 | 1.03  (0.92-1.15) | 637 | 1.18  (1.05-1.31) | 0.037 | 0.090 | 0.004 | 0.009 |
| Main model | 1.01  (0.97-1.04) | 568 | 1.00 (ref) | 534 | 0.99  (0.88-1.12) | 514 | 1.00  (0.89-1.13) | 468 | 0.98  (0.86-1.11) | 492 | 1.10  (0.97-1.24) | 0.402 | 0.636 | 0.153 | 0.265 |
| Additionally adjusted^c^ | 1.01  (0.97-1.04) | 568 | 1.00 (ref) | 534 | 0.99  (0.88-1.12) | 514 | 1.00  (0.89-1.13) | 468 | 0.98  (0.87-1.11) | 492 | 1.11  (0.98-1.25) | 0.361 | -- | 0.117 | -- |
| **Lung** |  |  |  |  |  |  |  |  |  |  |  |  |  |  |  |
| Simply adjusted model | 1.02  (1.01-1.03) | 940 | 1.00 (ref) | 966 | 1.10  (1.01-1.21) | 920 | 1.12  (1.02-1.22) | 920 | 1.20  (1.10-1.32) | 995 | 1.42  (1.29-1.55) | <0.001 | <0.001 | <0.001 | <0.001 |
| Main model | 1.01  (0.99-1.03) | 743 | 1.00 (ref) | 740 | 1.04  (0.94-1.15) | 705 | 1.03  (0.92-1.14) | 703 | 1.08  (0.97-1.20) | 751 | 1.19  (1.07-1.32) | 0.012 | 0.228 | 0.001 | 0.011 |
| Additionally adjusted^d^ | 1.01  (0.99-1.03) | 721 | 1.00 (ref) | 715 | 1.04  (0.93-1.15) | 682 | 1.03  (0.92-1.14) | 680 | 1.08  (0.97-1.20) | 726 | 1.20  (1.08-1.34) | 0.008 | -- | <0.001 | -- |
| **Malignant melanoma** |  |  |  |  |  |  |  |  |  |  |  |  |  |  |  |
| Simply adjusted model | 0.99  (0.94-1.05) | 885 | 1.00 (ref) | 862 | 1.02  (0.93-1.12) | 806 | 0.99  (0.90-1.09) | 730 | 0.95  (0.86-1.05) | 733 | 1.01  (0.91-1.11) | 0.652 | 0.652 | 0.748 | 0.748 |
| Main model | 0.98  (0.89-1.07) | 703 | 1.00 (ref) | 685 | 1.02  (0.92-1.13) | 637 | 0.98  (0.88-1.09) | 577 | 0.94  (0.84-1.05) | 570 | 0.99  (0.88-1.11) | 0.723 | 0.774 | 0.545 | 0.575 |
| Additionally adjusted^e^ | 0.97  (0.88-1.07) | 703 | 1.00 (ref) | 685 | 1.01  (0.91-1.13) | 637 | 0.97  (0.87-1.08) | 577 | 0.93  (0.84-1.05) | 570 | 0.98  (0.87-1.09) | 0.673 | -- | 0.411 | -- |
| **Connective soft tissue** |  |  |  |  |  |  |  |  |  |  |  |  |  |  |  |
| Simply adjusted model | 1.01  (0.99-1.04) | 591 | 1.00 (ref) | 582 | 1.04  (0.93-1.17) | 559 | 1.05  (0.94-1.18) | 512 | 1.02  (0.91-1.15) | 557 | 1.19  (1.06-1.34) | 0.043 | 0.091 | 0.006 | 0.012 |
| Main model | 1.01  (0.97-1.04) | 467 | 1.00 (ref) | 466 | 1.05  (0.92-1.19) | 430 | 1.00  (0.88-1.14) | 399 | 0.99  (0.86-1.13) | 426 | 1.12  (0.98-1.28) | 0.361 | 0.636 | 0.165 | 0.265 |
| **Breast** |  |  |  |  |  |  |  |  |  |  |  |  |  |  |  |
| Simply adjusted model | 1.00  (0.98-1.02) | 1,595 | 1.00 (ref) | 1,492 | 1.01  (0.94-1.08) | 1,436 | 1.04  (0.97-1.12) | 1,343 | 1.03  (0.96-1.11) | 1,315 | 1.10  (1.02-1.18) | 0.146 | 0.198 | 0.013 | 0.019 |
| Main model | 1.00  (0.96-1.03) | 1,247 | 1.00 (ref) | 1,170 | 1.01  (0.93-1.09) | 1,096 | 1.00  (0.92-1.09) | 1,034 | 1.01  (0.93-1.10) | 998 | 1.06  (0.97-1.15) | 0.706 | 0.774 | 0.195 | 0.267 |
| Additionally adjusted^f^ | 0.99  (0.96-1.03) | 841 | 1.00 (ref) | 780 | 1.00  (0.91-1.10) | 743 | 1.02  (0.93-1.13) | 711 | 1.04  (0.94-1.15) | 672 | 1.08  (0.97-1.20) | 0.562 | -- | 0.092 | -- |
| **Uterus** |  |  |  |  |  |  |  |  |  |  |  |  |  |  |  |
| Simply adjusted model | 1.01  (0.99-1.03) | 714 | 1.00 (ref) | 748 | 1.13  (1.02-1.25) | 681 | 1.10  (0.99-1.22) | 656 | 1.13  (1.02-1.26) | 646 | 1.22  (1.09-1.36) | 0.011 | 0.052 | 0.001 | 0.006 |
| Main model | 1.00  (0.97-1.04) | 570 | 1.00 (ref) | 587 | 1.09  (0.97-1.22) | 521 | 1.02  (0.90-1.15) | 502 | 1.05  (0.93-1.19) | 492 | 1.13  (0.99-1.27) | 0.325 | 0.636 | 0.116 | 0.265 |
| Additionally adjusted^g^ | 0.97  (0.82-1.14) | 308 | 1.00 (ref) | 315 | 1.12  (0.95-1.30) | 273 | 1.07  (0.91-1.26) | 252 | 1.06  (0.90-1.26) | 209 | 1.03  (0.86-1.23) | 0.713 | -- | 0.998 | -- |
| **Ovary** |  |  |  |  |  |  |  |  |  |  |  |  |  |  |  |
| Simply adjusted model | 1.01  (0.99-1.03) | 698 | 1.00 (ref) | 667 | 1.03  (0.92-1.14) | 624 | 1.02  (0.91-1.14) | 582 | 1.02  (0.91-1.14) | 615 | 1.16  (1.04-1.30) | 0.066 | 0.104 | 0.010 | 0.016 |
| Main model | 1.01  (0.97-1.04) | 548 | 1.00 (ref) | 524 | 1.01  (0.90-1.14) | 475 | 0.97  (0.85-1.09) | 450 | 0.98  (0.86-1.11) | 471 | 1.10  (0.97-1.24) | 0.342 | 0.636 | 0.197 | 0.267 |
| Additionally adjusted^g^ | 0.99  (0.92-1.07) | 277 | 1.00 (ref) | 248 | 1.00  (0.84-1.18) | 229 | 1.01  (0.85-1.21) | 196 | 0.95  (0.79-1.14) | 186 | 1.02  (0.85-1.24) | 0.954 | -- | 0.939 | -- |
| **Prostate** |  |  |  |  |  |  |  |  |  |  |  |  |  |  |  |
| Simply adjusted model | 0.95  (0.89-1.02) | 1,771 | 1.00 (ref) | 1,737 | 0.99  (0.93-1.06) | 1,624 | 0.93  (0.87-1.00) | 1,589 | 0.95  (0.89-1.02) | 1,550 | 0.96  (0.89-1.03) | 0.222 | 0.264 | 0.173 | 0.183 |
| Main model | 0.93  (0.86-1.00) | 1,468 | 1.00 (ref) | 1,453 | 1.01  (0.94-1.08) | 1,316 | 0.92  (0.85-0.99) | 1,301 | 0.96  (0.89-1.03) | 1,240 | 0.95  (0.88-1.03) | 0.099 | 0.594 | 0.148 | 0.265 |
| Additionally adjusted^h^ | 0.93  (0.86-1.01) | 1,428 | 1.00 (ref) | 1,421 | 1.02  (0.94-1.09) | 1,287 | 0.93  (0.86-1.00) | 1,273 | 0.97  (0.90-1.04) | 1,208 | 0.96  (0.89-1.04) | 0.129 | -- | 0.209 | -- |
| **Kidney** |  |  |  |  |  |  |  |  |  |  |  |  |  |  |  |
| Simply adjusted model | 1.01  (0.99-1.03) | 714 | 1.00 (ref) | 706 | 1.04  (0.94-1.16) | 649 | 1.01  (0.90-1.12) | 647 | 1.06  (0.95-1.18) | 660 | 1.16  (1.04-1.29) | 0.065 | 0.104 | 0.007 | 0.012 |
| Main model | 1.01  (0.98-1.04) | 571 | 1.00 (ref) | 552 | 1.01  (0.90-1.13) | 501 | 0.95  (0.84-1.07) | 506 | 1.02  (0.90-1.15) | 505 | 1.08  (0.96-1.22) | 0.374 | 0.636 | 0.167 | 0.265 |
| **Bladder** |  |  |  |  |  |  |  |  |  |  |  |  |  |  |  |
| Simply adjusted model | 1.01  (0.99-1.03) | 689 | 1.00 (ref) | 680 | 1.05  (0.94-1.16) | 639 | 1.04  (0.93-1.15) | 615 | 1.06  (0.95-1.19) | 640 | 1.19  (1.06-1.33) | 0.033 | 0.090 | 0.002 | 0.006 |
| Main model | 1.01  (0.97-1.04) | 557 | 1.00 (ref) | 541 | 1.02  (0.91-1.15) | 496 | 0.97  (0.86-1.10) | 478 | 1.00  (0.88-1.13) | 500 | 1.11  (0.98-1.26) | 0.266 | 0.636 | 0.096 | 0.265 |
| **Brain** |  |  |  |  |  |  |  |  |  |  |  |  |  |  |  |
| Simply adjusted model | 1.01  (0.98-1.04) | 656 | 1.00 (ref) | 637 | 1.03  (0.92-1.14) | 616 | 1.04  (0.93-1.16) | 562 | 1.01  (0.90-1.13) | 604 | 1.16  (1.03-1.29) | 0.094 | 0.137 | 0.019 | 0.025 |
| Main model | 1.00  (0.96-1.05) | 525 | 1.00 (ref) | 509 | 1.02  (0.90-1.15) | 475 | 0.98  (0.87-1.11) | 435 | 0.96  (0.84-1.09) | 459 | 1.07  (0.94-1.22) | 0.555 | 0.769 | 0.445 | 0.497 |
| **Thyroid** |  |  |  |  |  |  |  |  |  |  |  |  |  |  |  |
| Simply adjusted model | 1.01  (0.99-1.04) | 625 | 1.00 (ref) | 592 | 1.00  (0.90-1.12) | 580 | 1.03  (0.92-1.16) | 541 | 1.02  (0.91-1.15) | 566 | 1.14  (1.01-1.28) | 0.175 | 0.222 | 0.023 | 0.029 |
| Main model | 1.00  (0.97-1.04) | 498 | 1.00 (ref) | 470 | 0.99  (0.87-1.12) | 447 | 0.98  (0.86-1.11) | 417 | 0.97  (0.85-1.11) | 433 | 1.07  (0.94-1.22) | 0.644 | 0.774 | 0.331 | 0.393 |
| **Lymphoid and Hematopoietic Tissues** |  |  |  |  |  |  |  |  |  |  |  |  |  |  |  |
| Simply adjusted model | 1.02  (1.01-1.03) | 1,041 | 1.00 (ref) | 1,030 | 1.05  (0.96-1.14) | 982 | 1.05  (0.96-1.15) | 937 | 1.07  (0.98-1.17) | 938 | 1.15  (1.05-1.26) | 0.051 | 0.097 | 0.003 | 0.006 |
| Main model | 1.02  (1.00-1.03) | 819 | 1.00 (ref) | 817 | 1.05  (0.96-1.16) | 766 | 1.03  (0.93-1.14) | 715 | 1.03  (0.93-1.14) | 723 | 1.12  (1.01-1.24) | 0.287 | 0.636 | 0.057 | 0.265 |

Abbreviations: SD, standard deviation; CI, confidence interval; HR, hazards ratio; ref, reference.

The results from simply adjusted models revealed the associations of plasma omega-6/omega-3 ratio with cancer risk stratified by age and sex in general cohort. The main models were adjusted for general covariates including ethnicity (classified into White, Black, Asian, Others), Townsend deprivation index (continuous), assessment Center, BMI (kg/m2; continuous), smoking status (categorized as never, previous, current), alcohol intake status (categorized as never, previous, current), and physical activity (classified as low, moderate, high). The additionally adjusted models were adjusted for extra covariates for some specific types of cancer.

^a^ Additionally adjusted for gastroesophageal reflux disease at baseline and waist-hip ratio.

^b^ Additionally adjusted for diabetes at baseline, aspirin use, processed meat intake, waist-hip ratio, and family history.

^c^ Additionally adjusted for diabetes at baseline.

^d^ Additionally adjusted for family history.

^e^ Additionally adjusted for skin color, ease of skin tanning, use of sun/UV protection, childhood sunburn occasions, frequency of solarium/sunlamp use.

^f^ Restricted to female, and additionally adjusted for age when menarche started, hormone replacement therapy use, oral contraceptive use, number of live births, menopausal status, hysterectomy status, and family history.

^g^ Restricted to female, and additionally adjusted for age when menarche started, hormone replacement therapy use, oral contraceptive use, number of live births, menopausal status, hysterectomy status.

^h^ Restricted to male, and additionally adjusted for family history.

^i^ Used likelihood ratio test to compare the full model with reduced model.

^j^ Based on False Discovery Rate (FDR) to calculate the adjusted p-values for simply adjusted models and main models among 19 cancer sites.

^k^ Used the median value of each quintile as a continuous variable within the models.

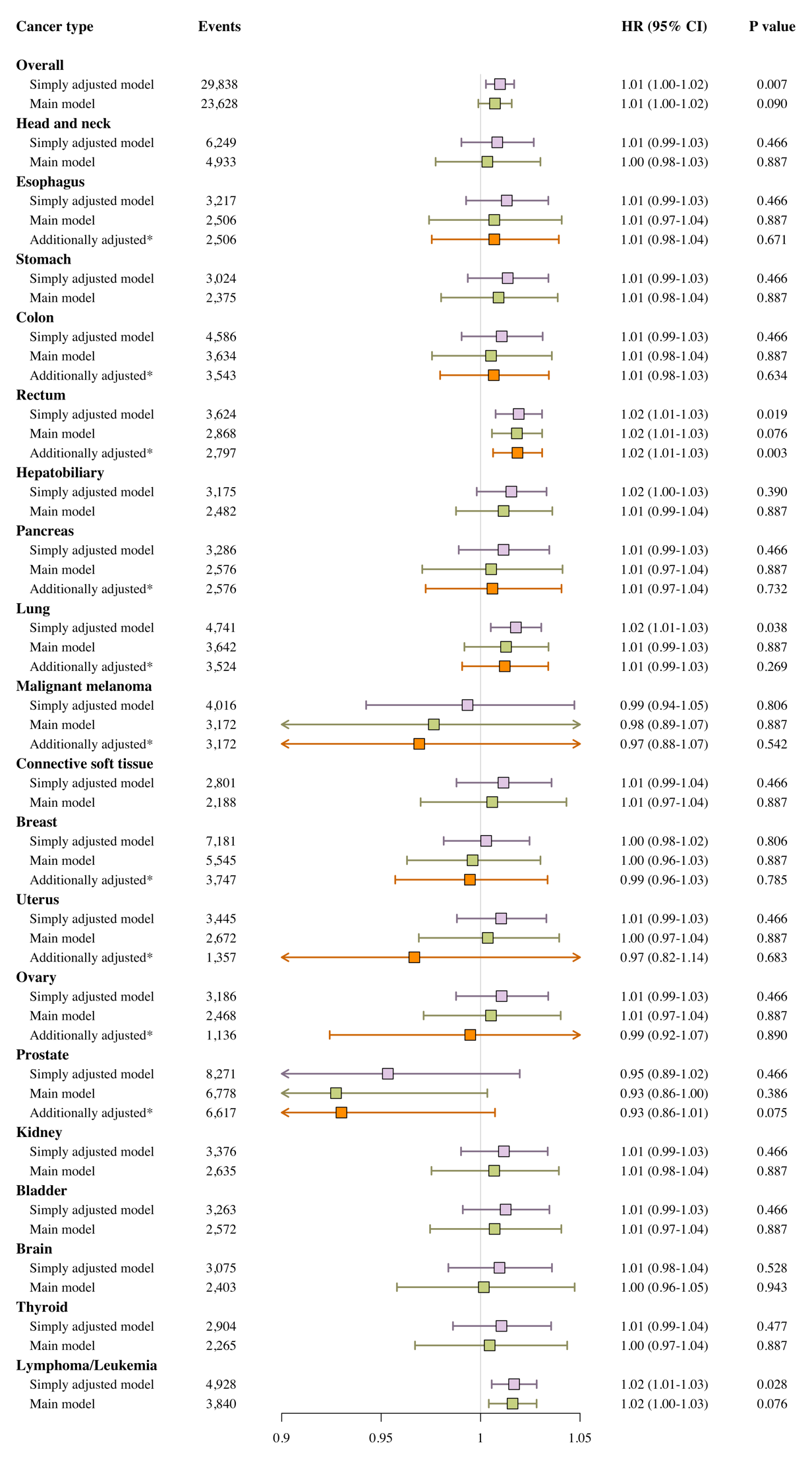

**Figure S1. Risk estimates of incidence of overall cancer and 19 cancer sites for 1-SD increase of plasma omega-6/omega-3 ratio.** P values for simply adjusted and main models were corrected for the multiple testing of 19 site-specific cancers. The results from simply adjusted models revealed the associations stratified by age and sex in general cohort. The main models were adjusted for general covariates including ethnicity (classified into White, Black, Asian, Others), Townsend deprivation index (continuous), assessment Center, BMI (kg/m2; continuous), smoking status (categorized as never, previous, current), alcohol intake status (categorized as never, previous, current), and physical activity (classified as low, moderate, high).

* For esophagus cancer, additionally adjusted for gastroesophageal reflux disease at baseline and waist-hip ratio. For colon cancer and rectum cancer, additionally adjusted for diabetes at baseline, aspirin use, processed meat intake, waist-hip ratio, and family history. For pancreas cancer, additionally adjusted for diabetes at baseline. For lung cancer, additionally adjusted for family history. For malignant melanoma cancer, additionally adjusted for skin color, ease of skin tanning, use of sun/UV protection, childhood sunburn occasions, frequency of solarium/sunlamp use. For breast cancer, restricted to female, and additionally adjusted for age when menarche started, hormone replacement therapy use, oral contraceptive use, number of live births, menopausal status, hysterectomy status, and family history. For uterus and ovary cancer, restricted to female, and additionally adjusted for age when menarche started, hormone replacement therapy use, oral contraceptive use, number of live births, menopausal status, hysterectomy status. For prostate cancer, restricted to male, and additionally adjusted for family history.

Table S4. Associations^a^ of plasma omega-6% with incidence of overall cancer in the UK Biobank, additionally adjusted for omega-3%

| Omega-6  Variable forms | **Overall Cancer** | | | | | | |
| --- | --- | --- | --- | --- | --- | --- | --- |
|  | Simply adjusted model^b^ | | |  | Main model^c^ | | |
|  | Events | HR  (95% CI) | P values |  | Events | HR  (95% CI) | P values |
| Continuous (per 1-SD) | 29,838 | 0.94  (0.93-0.95) | < 0.001 |  | 23,628 | 0.97  (0.96-0.99) | < 0.001 |
| Quintiles (median) |  |  | | | | | |
| 1 (32.9) | 6,787 | 1.00  (ref) | -- |  | 5,356 | 1.00  (ref) | -- |
| 2 (36.4) | 6,427 | 0.96  (0.93-0.99) | 0.016 |  | 5,064 | 0.99  (0.95-1.03) | 0.559 |
| 3 (38.4) | 6,118 | 0.94  (0.90-0.97) | < 0.001 |  | 4,804 | 0.98  (0.94-1.02) | 0.339 |
| 4 (40.0) | 5,579 | 0.88  (0.85-0.91) | < 0.001 |  | 4,420 | 0.95  (0.91-0.99) | 0.012 |
| 5 (42.1) | 4,927 | 0.83  (0.80-0.86) | < 0.001 |  | 3,984 | 0.93  (0.89-0.97) | 0.001 |
| *P for overall* | *< 0.001* | | |  | *0.004* | | |
| *P for trend* | *< 0.001* | | |  | *< 0.001* | | |

Abbreviations: omega-6%, omega-6 fatty acids to total fatty acids percentage; omega-3%, omega-3 fatty acids to total fatty acids percentage; SD, standard deviation; CI, confidence interval; HR, hazards ratio; ref, reference.

^a^ From Cox proportional hazards regression.

^b^ Stratified by age and sex, additionally adjusted for omega-3%.

^c^ Additionally adjusted for omega-3%, ethnicity (classified into White, Black, Asian, Others), Townsend deprivation index (continuous), assessment Center, BMI (kg/m2; continuous), smoking status (categorized as never, previous, current), alcohol intake status (categorized as never, previous, current), and physical activity (classified as low, moderate, high).

Table S5. Associations^a^ of plasma omega-3% with incidence of overall cancer in the UK Biobank, additionally adjusted for omega-6%

| Omega-3  Variable forms | **Overall Cancer** | | | | | | |
| --- | --- | --- | --- | --- | --- | --- | --- |
|  | Simply adjusted model^b^ | | |  | Main model^c^ | | |
|  | Events | HR  (95% CI) | P values |  | Events | HR  (95% CI) | P values |
| Continuous (per 1-SD) | 29,838 | 0.95  (0.94-0.96) | < 0.001 |  | 23,628 | 0.98  (0.97-1.00) | 0.008 |
| Quintiles (median) |  |  | | | | | |
| 1 (2.7) | 5,864 | 1.00  (ref) | -- |  | 4,546 | 1.00  (ref) | -- |
| 2 (3.5) | 5,837 | 0.94  (0.91-0.97) | 0.001 |  | 4,603 | 0.96  (0.93-1.01) | 0.088 |
| 3 (4.2) | 5,935 | 0.91  (0.88-0.94) | < 0.001 |  | 4,688 | 0.94  (0.91-0.98) | 0.006 |
| 4 (4.9) | 6,086 | 0.90  (0.87-0.93) | < 0.001 |  | 4,838 | 0.95  (0.91-0.98) | 0.007 |
| 5 (6.3) | 6,116 | 0.86  (0.83-0.90) | < 0.001 |  | 4,953 | 0.94  (0.90-0.98) | 0.004 |
| *P for overall* | *< 0.001* | | |  | *0.023* | | |
| *P for trend* | *< 0.001* | | |  | *0.005* | | |

Abbreviations: omega-3%, omega-3 fatty acids to total fatty acids percentage; omega-6%, omega-6 fatty acids to total fatty acids percentage; SD, standard deviation; CI, confidence interval; HR, hazards ratio; ref, reference.

^a^ From Cox proportional hazards regression.

^b^ Stratified by age and sex, additionally adjusted for omega-6%.

^c^ Additionally adjusted for omega-6%, ethnicity (classified into White, Black, Asian, Others), Townsend deprivation index (continuous), assessment Center, BMI (kg/m2; continuous), smoking status (categorized as never, previous, current), alcohol intake status (categorized as never, previous, current), and physical activity (classified as low, moderate, high).

Table S6. Associations of the plasma DHA% with the incidence of overall cancer and 19 cancer sites in the UK Biobank

| **Cancer Type** | **Per 1-SD** | **Quintiles** | | | | | | | | | | **P for overall**^i^ | **Adjusted P for overall**^j^ | **P for trend**^k^ | **Adjusted P for trend**^j^ |
| --- | --- | --- | --- | --- | --- | --- | --- | --- | --- | --- | --- | --- | --- | --- | --- |
|  | HR  (95% CI) | **1** | | **2** | | **3** | | **4** | | **5** | |  |  |  |  |
|  |  | Events | HR  (95% CI) | Events | HR  (95% CI) | Events | HR  (95% CI) | Events | HR  (95% CI) | Events | HR  (95% CI) |  |  |  |  |
| **Overall** |  |  |  |  |  |  |  |  |  |  |  |  |  |  |  |
| Simply adjusted model | 0.95 (0.94-0.96) | 6,226 | 1.00 (ref) | 6,026 | 0.95 (0.91-0.98) | 5,774 | 0.90 (0.87-0.93) | 5,887 | 0.90 (0.86-0.93) | 5,925 | 0.87 (0.84-0.90) | < 0.001 | -- | < 0.001 | -- |
| Main model | 0.99 (0.97-1.00) | 4,824 | 1.00 (ref) | 4,707 | 0.98 (0.94-1.02) | 4,571 | 0.95 (0.91-0.99) | 4,711 | 0.97 (0.93-1.01) | 4,815 | 0.97 (0.93-1.01) | 0.206 | -- | 0.144 | -- |
| **Head and neck** |  |  |  |  |  |  |  |  |  |  |  |  |  |  |  |
| Simply adjusted model | 0.94 (0.92-0.97) | 1,317 | 1.00 (ref) | 1,246 | 0.91 (0.84-0.98) | 1,237 | 0.88 (0.81-0.95) | 1,176 | 0.81 (0.75-0.88) | 1,273 | 0.85 (0.79-0.92) | < 0.001 | < 0.001 | < 0.001 | < 0.001 |
| Main model | 0.98 (0.95-1.00) | 1,022 | 1.00 (ref) | 976 | 0.94 (0.86-1.02) | 965 | 0.92 (0.84-1.01) | 938 | 0.88 (0.80-0.96) | 1,032 | 0.94 (0.86-1.03) | 0.079 | 0.151 | 0.163 | 0.182 |
| **Esophagus** |  |  |  |  |  |  |  |  |  |  |  |  |  |  |  |
| Simply adjusted model | 0.89 (0.86-0.92) | 762 | 1.00 (ref) | 658 | 0.85 (0.77-0.94) | 641 | 0.82 (0.74-0.91) | 557 | 0.70 (0.62-0.78) | 599 | 0.72 (0.64-0.80) | < 0.001 | < 0.001 | < 0.001 | < 0.001 |
| Main model | 0.94 (0.90-0.98) | 574 | 1.00 (ref) | 519 | 0.92 (0.82-1.04) | 490 | 0.89 (0.78-1.00) | 447 | 0.81 (0.71-0.92) | 476 | 0.84 (0.74-0.95) | 0.011 | 0.052 | 0.002 | 0.008 |
| Additionally adjusted^a^ | 0.95  (0.91-0.99) | 574 | 1.00 (ref) | 519 | 0.93  (0.83-1.05) | 490 | 0.90  (0.80-1.02) | 447 | 0.83  (0.73-0.94) | 476 | 0.87  (0.76-0.99) | 0.051 | -- | 0.012 | -- |
| **Stomach** |  |  |  |  |  |  |  |  |  |  |  |  |  |  |  |
| Simply adjusted model | 0.89 (0.86-0.92) | 688 | 1.00 (ref) | 640 | 0.91 (0.82-1.01) | 592 | 0.83 (0.74-0.93) | 540 | 0.74 (0.66-0.83) | 564 | 0.74 (0.66-0.83) | < 0.001 | < 0.001 | < 0.001 | < 0.001 |
| Main model | 0.93 (0.89-0.97) | 526 | 1.00 (ref) | 508 | 0.98 (0.87-1.11) | 458 | 0.89 (0.79-1.02) | 434 | 0.84 (0.73-0.96) | 449 | 0.84 (0.74-0.97) | 0.022 | 0.084 | 0.002 | 0.008 |
| **Colon** |  |  |  |  |  |  |  |  |  |  |  |  |  |  |  |
| Simply adjusted model | 0.91 (0.88-0.94) | 1,033 | 1.00 (ref) | 941 | 0.89 (0.81-0.97) | 872 | 0.81 (0.74-0.88) | 851 | 0.76 (0.70-0.84) | 889 | 0.76 (0.69-0.83) | < 0.001 | < 0.001 | < 0.001 | < 0.001 |
| Main model | 0.94 (0.91-0.97) | 798 | 1.00 (ref) | 761 | 0.95 (0.86-1.05) | 688 | 0.86 (0.78-0.96) | 678 | 0.84 (0.75-0.93) | 709 | 0.85 (0.76-0.94) | 0.002 | 0.013 | < 0.001 | < 0.001 |
| Additionally adjusted^b^ | 0.94  (0.91-0.97) | 780 | 1.00 (ref) | 733 | 0.94  (0.85-1.04) | 674 | 0.86  (0.77-0.96) | 664 | 0.84  (0.75-0.93) | 692 | 0.84  (0.76-0.94) | 0.004 | -- | 0.001 | -- |
| **Rectum** |  |  |  |  |  |  |  |  |  |  |  |  |  |  |  |
| Simply adjusted model | 0.92 (0.89-0.95) | 784 | 1.00 (ref) | 759 | 0.96 (0.87-1.06) | 728 | 0.91 (0.83-1.01) | 673 | 0.83 (0.75-0.92) | 680 | 0.81 (0.73-0.90) | < 0.001 | < 0.001 | < 0.001 | < 0.001 |
| Main model | 0.96 (0.92-0.99) | 600 | 1.00 (ref) | 607 | 1.02 (0.91-1.15) | 569 | 0.97 (0.87-1.09) | 545 | 0.92 (0.82-1.04) | 547 | 0.91 (0.80-1.02) | 0.221 | 0.233 | 0.035 | 0.048 |
| Additionally adjusted^b^ | 0.95  (0.92-0.99) | 584 | 1.00 (ref) | 587 | 1.01  (0.90-1.14) | 556 | 0.96  (0.85-1.08) | 535 | 0.92  (0.81-1.04) | 535 | 0.90  (0.79-1.01) | 0.239 | -- | 0.030 | -- |
| **Hepatobiliary** |  |  |  |  |  |  |  |  |  |  |  |  |  |  |  |
| Simply adjusted model | 0.88 (0.84-0.91) | 750 | 1.00 (ref) | 677 | 0.88 (0.80-0.98) | 607 | 0.78 (0.70-0.87) | 556 | 0.70 (0.62-0.78) | 585 | 0.70 (0.63-0.78) | < 0.001 | < 0.001 | < 0.001 | < 0.001 |
| Main model | 0.93 (0.89-0.97) | 563 | 1.00 (ref) | 538 | 0.97 (0.86-1.10) | 472 | 0.87 (0.76-0.98) | 447 | 0.81 (0.71-0.92) | 462 | 0.82 (0.72-0.93) | 0.002 | 0.013 | < 0.001 | < 0.001 |
| **Pancreas** |  |  |  |  |  |  |  |  |  |  |  |  |  |  |  |
| Simply adjusted model | 0.90 (0.87-0.93) | 728 | 1.00 (ref) | 688 | 0.92 (0.82-1.02) | 647 | 0.84 (0.76-0.94) | 584 | 0.74 (0.66-0.82) | 639 | 0.77 (0.69-0.86) | < 0.001 | < 0.001 | < 0.001 | < 0.001 |
| Main model | 0.95 (0.91-0.99) | 555 | 1.00 (ref) | 546 | 0.99 (0.88-1.11) | 492 | 0.90 (0.79-1.02) | 471 | 0.85 (0.75-0.96) | 512 | 0.89 (0.78-1.01) | 0.045 | 0.122 | 0.018 | 0.031 |
| Additionally adjusted^c^ | 0.94  (0.91-0.98) | 555 | 1.00 (ref) | 546 | 0.99  (0.88-1.11) | 492 | 0.89  (0.79-1.01) | 471 | 0.84  (0.74-0.96) | 512 | 0.89  (0.78-1.01) | 0.035 | -- | 0.014 | -- |
| **Lung** |  |  |  |  |  |  |  |  |  |  |  |  |  |  |  |
| Simply adjusted model | 0.83 (0.81-0.86) | 1,163 | 1.00 (ref) | 1,036 | 0.86 (0.79-0.93) | 924 | 0.75 (0.69-0.82) | 801 | 0.63 (0.57-0.69) | 817 | 0.60 (0.55-0.66) | < 0.001 | < 0.001 | < 0.001 | < 0.001 |
| Main model | 0.92 (0.88-0.95) | 862 | 1.00 (ref) | 781 | 0.94 (0.85-1.03) | 710 | 0.88 (0.79-0.97) | 634 | 0.79 (0.71-0.88) | 655 | 0.80 (0.72-0.89) | < 0.001 | < 0.001 | < 0.001 | < 0.001 |
| Additionally adjusted^d^ | 0.92  (0.89-0.95) | 827 | 1.00 (ref) | 743 | 0.93  (0.84-1.03) | 698 | 0.89  (0.80-0.98) | 617 | 0.79  (0.71-0.88) | 639 | 0.80  (0.72-0.90) | < 0.001 | -- | < 0.001 | -- |
| **Malignant melanoma** |  |  |  |  |  |  |  |  |  |  |  |  |  |  |  |
| Simply adjusted model | 0.97 (0.94-1.00) | 816 | 1.00 (ref) | 791 | 0.95 (0.86-1.04) | 789 | 0.93 (0.84-1.03) | 793 | 0.91 (0.83-1.01) | 827 | 0.92 (0.83-1.02) | 0.412 | 0.412 | 0.095 | 0.095 |
| Main model | 0.99 (0.96-1.03) | 625 | 1.00 (ref) | 637 | 1.01 (0.90-1.13) | 612 | 0.97 (0.86-1.08) | 643 | 0.99 (0.89-1.11) | 655 | 0.99 (0.88-1.11) | 0.956 | 0.956 | 0.772 | 0.772 |
| Additionally adjusted^e^ | 1.00  (0.96-1.03) | 625 | 1.00 (ref) | 637 | 1.01  (0.90-1.13) | 612 | 0.97  (0.87-1.09) | 643 | 1.00  (0.89-1.12) | 655 | 1.00  (0.89-1.12) | 0.961 | -- | 0.996 | -- |
| **Connective soft tissue** |  |  |  |  |  |  |  |  |  |  |  |  |  |  |  |
| Simply adjusted model | 0.90 (0.87-0.94) | 626 | 1.00 (ref) | 582 | 0.90 (0.81-1.01) | 559 | 0.86 (0.76-0.96) | 502 | 0.75 (0.66-0.84) | 532 | 0.76 (0.67-0.85) | < 0.001 | < 0.001 | < 0.001 | < 0.001 |
| Main model | 0.94 (0.90-0.98) | 474 | 1.00 (ref) | 461 | 0.98 (0.86-1.11) | 428 | 0.91 (0.80-1.04) | 403 | 0.85 (0.74-0.97) | 422 | 0.86 (0.75-0.99) | 0.078 | 0.151 | 0.011 | 0.025 |
| **Breast** |  |  |  |  |  |  |  |  |  |  |  |  |  |  |  |
| Simply adjusted model | 0.94 (0.92-0.96) | 1,217 | 1.00 (ref) | 1,416 | 0.97 (0.90-1.05) | 1,406 | 0.87 (0.81-0.94) | 1,528 | 0.87 (0.81-0.94) | 1,614 | 0.86 (0.80-0.93) | < 0.001 | < 0.001 | < 0.001 | < 0.001 |
| Main model | 0.97 (0.94-1.00) | 904 | 1.00 (ref) | 1,087 | 1.02 (0.93-1.11) | 1,063 | 0.91 (0.83-1.00) | 1,204 | 0.95 (0.87-1.04) | 1,287 | 0.96 (0.88-1.05) | 0.104 | 0.151 | 0.235 | 0.248 |
| Additionally adjusted^f^ | 0.99  (0.95-1.02) | 491 | 1.00 (ref) | 684 | 1.02  (0.91-1.14) | 718 | 0.91  (0.81-1.03) | 881 | 1.00  (0.89-1.12) | 973 | 1.00  (0.89-1.12) | 0.241 | -- | 0.839 | -- |
| **Uterus** |  |  |  |  |  |  |  |  |  |  |  |  |  |  |  |
| Simply adjusted model | 0.88 (0.85-0.91) | 739 | 1.00 (ref) | 713 | 0.89 (0.80-0.99) | 693 | 0.82 (0.74-0.92) | 640 | 0.72 (0.65-0.80) | 660 | 0.70 (0.63-0.78) | < 0.001 | < 0.001 | < 0.001 | < 0.001 |
| Main model | 0.94 (0.90-0.98) | 553 | 1.00 (ref) | 550 | 0.97 (0.86-1.09) | 532 | 0.93 (0.82-1.05) | 508 | 0.86 (0.76-0.98) | 529 | 0.87 (0.77-0.99) | 0.097 | 0.151 | 0.011 | 0.025 |
| Additionally adjusted^g^ | 0.98  (0.92-1.03) | 202 | 1.00 (ref) | 246 | 0.96  (0.80-1.15) | 276 | 0.93  (0.77-1.11) | 300 | 0.94  (0.78-1.13) | 333 | 0.95  (0.79-1.14) | 0.953 | -- | 0.681 | -- |
| **Ovary** |  |  |  |  |  |  |  |  |  |  |  |  |  |  |  |
| Simply adjusted model | 0.91 (0.88-0.94) | 677 | 1.00 (ref) | 654 | 0.91 (0.81-1.01) | 618 | 0.82 (0.74-0.92) | 600 | 0.76 (0.68-0.85) | 637 | 0.77 (0.69-0.85) | < 0.001 | < 0.001 | < 0.001 | < 0.001 |
| Main model | 0.95 (0.91-0.99) | 517 | 1.00 (ref) | 506 | 0.95 (0.84-1.07) | 469 | 0.86 (0.76-0.98) | 475 | 0.84 (0.74-0.96) | 501 | 0.86 (0.75-0.97) | 0.045 | 0.122 | 0.009 | 0.025 |
| Additionally adjusted^g^ | 0.99  (0.93-1.05) | 167 | 1.00 (ref) | 197 | 0.88  (0.72-1.08) | 206 | 0.78  (0.64-0.96) | 265 | 0.90  (0.74-1.09) | 301 | 0.9  (0.74-1.10) | 0.209 | -- | 0.741 | -- |
| **Prostate** |  |  |  |  |  |  |  |  |  |  |  |  |  |  |  |
| Simply adjusted model | 1.02 (1.00-1.05) | 1,786 | 1.00 (ref) | 1,722 | 1.05 (0.98-1.12) | 1,632 | 1.07 (1.00-1.15) | 1,559 | 1.07 (1.00-1.14) | 1,572 | 1.09 (1.02-1.17) | 0.106 | 0.112 | 0.013 | 0.014 |
| Main model | 1.03 (1.01-1.06) | 1,434 | 1.00 (ref) | 1,407 | 1.06 (0.98-1.14) | 1,326 | 1.07 (0.99-1.15) | 1,300 | 1.08 (1.00-1.17) | 1,311 | 1.10 (1.02-1.19) | 0.133 | 0.168 | 0.014 | 0.027 |
| Additionally adjusted^h^ | 1.03  (1.01-1.05) | 1,390 | 1.00 (ref) | 1,378 | 1.07  (0.99-1.15) | 1,302 | 1.07  (0.99-1.16) | 1,268 | 1.08  (1.00-1.17) | 1,279 | 1.10  (1.02-1.19) | 0.154 | -- | 0.026 | -- |
| **Kidney** |  |  |  |  |  |  |  |  |  |  |  |  |  |  |  |
| Simply adjusted model | 0.91 (0.87-0.94) | 780 | 1.00 (ref) | 690 | 0.87 (0.79-0.96) | 661 | 0.83 (0.74-0.92) | 608 | 0.74 (0.67-0.83) | 637 | 0.75 (0.67-0.83) | < 0.001 | < 0.001 | < 0.001 | < 0.001 |
| Main model | 0.96 (0.92-1.00) | 585 | 1.00 (ref) | 544 | 0.95 (0.85-1.07) | 508 | 0.91 (0.80-1.02) | 487 | 0.87 (0.76-0.98) | 511 | 0.89 (0.79-1.01) | 0.181 | 0.202 | 0.039 | 0.049 |
| **Bladder** |  |  |  |  |  |  |  |  |  |  |  |  |  |  |  |
| Simply adjusted model | 0.90 (0.87-0.93) | 755 | 1.00 (ref) | 662 | 0.86 (0.78-0.96) | 643 | 0.83 (0.75-0.92) | 588 | 0.74 (0.66-0.83) | 615 | 0.74 (0.67-0.83) | < 0.001 | < 0.001 | < 0.001 | < 0.001 |
| Main model | 0.94 (0.91-0.98) | 577 | 1.00 (ref) | 524 | 0.92 (0.82-1.04) | 496 | 0.89 (0.79-1.00) | 482 | 0.86 (0.76-0.97) | 493 | 0.86 (0.75-0.97) | 0.092 | 0.151 | 0.012 | 0.025 |
| **Brain** |  |  |  |  |  |  |  |  |  |  |  |  |  |  |  |
| Simply adjusted model | 0.92 (0.88-0.95) | 678 | 1.00 (ref) | 643 | 0.93 (0.83-1.03) | 609 | 0.87 (0.78-0.97) | 543 | 0.76 (0.67-0.85) | 602 | 0.80 (0.72-0.90) | < 0.001 | < 0.001 | < 0.001 | < 0.001 |
| Main model | 0.96 (0.92-1.00) | 513 | 1.00 (ref) | 506 | 0.99 (0.88-1.12) | 465 | 0.92 (0.81-1.04) | 436 | 0.85 (0.74-0.97) | 483 | 0.92 (0.80-1.04) | 0.098 | 0.151 | 0.060 | 0.071 |
| **Thyroid** |  |  |  |  |  |  |  |  |  |  |  |  |  |  |  |
| Simply adjusted model | 0.91 (0.88-0.95) | 636 | 1.00 (ref) | 606 | 0.92 (0.82-1.03) | 573 | 0.85 (0.76-0.95) | 524 | 0.76 (0.67-0.85) | 565 | 0.78 (0.69-0.87) | < 0.001 | < 0.001 | < 0.001 | < 0.001 |
| Main model | 0.95 (0.91-0.99) | 479 | 1.00 (ref) | 478 | 0.99 (0.87-1.13) | 439 | 0.91 (0.80-1.04) | 419 | 0.86 (0.75-0.98) | 450 | 0.89 (0.78-1.02) | 0.111 | 0.151 | 0.032 | 0.048 |
| **Lymphoid and Hematopoietic Tissues** |  |  |  |  |  |  |  |  |  |  |  |  |  |  |  |
| Simply adjusted model | 0.93 (0.91-0.96) | 1,055 | 1.00 (ref) | 1,018 | 0.95 (0.87-1.03) | 973 | 0.89 (0.82-0.98) | 904 | 0.81 (0.74-0.89) | 978 | 0.84 (0.77-0.92) | < 0.001 | < 0.001 | < 0.001 | < 0.001 |
| Main model | 0.96 (0.93-1.00) | 799 | 1.00 (ref) | 799 | 1.00 (0.91-1.11) | 751 | 0.95 (0.86-1.05) | 726 | 0.90 (0.81-1.00) | 765 | 0.92 (0.83-1.02) | 0.162 | 0.192 | 0.035 | 0.048 |

Abbreviations: DHA%, DHA to total fatty acids percentage; SD, standard deviation; CI, confidence interval; HR, hazards ratio; ref, reference.

The results from simply adjusted models revealed the associations of plasma DHA% with cancer risk stratified by age and sex in general cohort. The main models were adjusted for general covariates including ethnicity (classified into White, Black, Asian, Others), Townsend deprivation index (continuous), assessment Center, BMI (kg/m2; continuous), smoking status (categorized as never, previous, current), alcohol intake status (categorized as never, previous, current), and physical activity (classified as low, moderate, high). The additionally adjusted models were adjusted for extra covariates for some specific types of cancer.

^a^ Additionally adjusted for gastroesophageal reflux disease at baseline and waist-hip ratio.

^b^ Additionally adjusted for diabetes at baseline, aspirin use, processed meat intake, waist-hip ratio, and family history.

^c^ Additionally adjusted for diabetes at baseline.

^d^ Additionally adjusted for family history.

^e^ Additionally adjusted for skin color, ease of skin tanning, use of sun/UV protection, childhood sunburn occasions, frequency of solarium/sunlamp use.

^f^ Restricted to female, and additionally adjusted for age when menarche started, hormone replacement therapy use, oral contraceptive use, number of live births, menopausal status, hysterectomy status, and family history.

^g^ Restricted to female, and additionally adjusted for age when menarche started, hormone replacement therapy use, oral contraceptive use, number of live births, menopausal status, hysterectomy status.

^h^ Restricted to male, and additionally adjusted for family history.

^i^ Used likelihood ratio test to compare the full model with reduced model.

^j^ Based on False Discovery Rate (FDR) to calculate the adjusted p-values for simply adjusted models and main models among 19 cancer sites.

^k^ Used the median value of each quintile as a continuous variable within the models.

Table S7. Associations of the plasma LA% with the incidence of overall cancer and 19 cancer sites in the UK Biobank

| **Cancer Type** | **Per 1-SD** | **Quintiles** | | | | | | | | | | **P for overall**^i^ | **Adjusted P for overall**^j^ | **P for trend**^k^ | **Adjusted P for trend**^j^ |
| --- | --- | --- | --- | --- | --- | --- | --- | --- | --- | --- | --- | --- | --- | --- | --- |
|  | HR  (95% CI) | **1** | | **2** | | **3** | | **4** | | **5** | |  |  |  |  |
|  |  | Events | HR  (95% CI) | Events | HR  (95% CI) | Events | HR  (95% CI) | Events | HR  (95% CI) | Events | HR  (95% CI) |  |  |  |  |
| **Overall** |  |  |  |  |  |  |  |  |  |  |  |  |  |  |  |
| Simply adjusted model | 0.93 (0.92-0.94) | 7,159 | 1.00 (ref) | 6,454 | 0.94 (0.91-0.98) | 5,848 | 0.90 (0.87-0.93) | 5,424 | 0.86 (0.83-0.89) | 4,953 | 0.81 (0.79-0.85) | < 0.001 | -- | < 0.001 | -- |
| Main model | 0.96 (0.95-0.97) | 5,635 | 1.00 (ref) | 5,098 | 0.97 (0.94-1.01) | 4,586 | 0.93 (0.89-0.97) | 4,299 | 0.92 (0.88-0.96) | 4,010 | 0.90 (0.86-0.94) | < 0.001 | -- | < 0.001 | -- |
| **Head and neck** |  |  |  |  |  |  |  |  |  |  |  |  |  |  |  |
| Simply adjusted model | 0.92 (0.90-0.95) | 1,444 | 1.00 (ref) | 1,335 | 0.94 (0.87-1.01) | 1,201 | 0.86 (0.80-0.93) | 1,207 | 0.88 (0.82-0.95) | 1,062 | 0.79 (0.73-0.86) | < 0.001 | < 0.001 | < 0.001 | < 0.001 |
| Main model | 0.95 (0.92-0.98) | 1,139 | 1.00 (ref) | 1,041 | 0.95 (0.87-1.04) | 935 | 0.88 (0.81-0.96) | 950 | 0.93 (0.85-1.01) | 868 | 0.87 (0.79-0.96) | 0.022 | 0.025 | 0.004 | 0.004 |
| **Esophagus** |  |  |  |  |  |  |  |  |  |  |  |  |  |  |  |
| Simply adjusted model | 0.87 (0.84-0.90) | 844 | 1.00 (ref) | 749 | 0.94 (0.85-1.03) | 605 | 0.80 (0.72-0.89) | 555 | 0.76 (0.69-0.85) | 464 | 0.67 (0.59-0.75) | < 0.001 | < 0.001 | < 0.001 | < 0.001 |
| Main model | 0.90 (0.86-0.94) | 652 | 1.00 (ref) | 591 | 1.00 (0.89-1.12) | 470 | 0.85 (0.76-0.96) | 431 | 0.84 (0.74-0.95) | 362 | 0.75 (0.65-0.86) | < 0.001 | < 0.001 | < 0.001 | < 0.001 |
| Additionally adjusted^a^ | 0.92  (0.88-0.96) | 652 | 1.00 (ref) | 591 | 1.01  (0.91-1.13) | 470 | 0.88  (0.78-0.99) | 431 | 0.87  (0.77-0.99) | 362 | 0.78  (0.68-0.9) | < 0.001 | -- | < 0.001 | -- |
| **Stomach** |  |  |  |  |  |  |  |  |  |  |  |  |  |  |  |
| Simply adjusted model | 0.89 (0.86-0.93) | 779 | 1.00 (ref) | 668 | 0.90 (0.81-1.00) | 566 | 0.80 (0.72-0.90) | 548 | 0.81 (0.73-0.91) | 463 | 0.71 (0.64-0.80) | < 0.001 | < 0.001 | < 0.001 | < 0.001 |
| Main model | 0.93 (0.89-0.97) | 601 | 1.00 (ref) | 537 | 0.98 (0.87-1.10) | 438 | 0.85 (0.75-0.97) | 435 | 0.91 (0.80-1.03) | 364 | 0.80 (0.69-0.92) | 0.007 | 0.010 | 0.001 | 0.001 |
| **Colon** |  |  |  |  |  |  |  |  |  |  |  |  |  |  |  |
| Simply adjusted model | 0.89 (0.87-0.92) | 1,172 | 1.00 (ref) | 1,030 | 0.92 (0.85-1.00) | 863 | 0.81 (0.74-0.89) | 796 | 0.78 (0.71-0.86) | 725 | 0.74 (0.68-0.82) | < 0.001 | < 0.001 | < 0.001 | < 0.001 |
| Main model | 0.93 (0.89-0.96) | 920 | 1.00 (ref) | 816 | 0.96 (0.88-1.06) | 691 | 0.87 (0.79-0.97) | 637 | 0.86 (0.78-0.96) | 570 | 0.82 (0.73-0.91) | 0.001 | 0.002 | < 0.001 | < 0.001 |
| Additionally adjusted^b^ | 0.94  (0.91-0.98) | 899 | 1.00 (ref) | 793 | 0.98  (0.89-1.08) | 672 | 0.90  (0.81-1.00) | 621 | 0.90  (0.81-1.00) | 558 | 0.86  (0.77-0.97) | 0.059 | -- | 0.004 | -- |
| **Rectum** |  |  |  |  |  |  |  |  |  |  |  |  |  |  |  |
| Simply adjusted model | 0.89 (0.86-0.92) | 967 | 1.00 (ref) | 780 | 0.85 (0.78-0.94) | 674 | 0.78 (0.70-0.86) | 638 | 0.77 (0.69-0.85) | 565 | 0.71 (0.64-0.79) | < 0.001 | < 0.001 | < 0.001 | < 0.001 |
| Main model | 0.92 (0.88-0.95) | 755 | 1.00 (ref) | 622 | 0.90 (0.80-1.00) | 535 | 0.82 (0.73-0.92) | 504 | 0.82 (0.73-0.92) | 452 | 0.77 (0.68-0.87) | < 0.001 | < 0.001 | < 0.001 | < 0.001 |
| Additionally adjusted^b^ | 0.93  (0.89-0.97) | 737 | 1.00 (ref) | 608 | 0.91  (0.82-1.02) | 523 | 0.84  (0.75-0.95) | 488 | 0.84  (0.75-0.95) | 441 | 0.8  (0.71-0.91) | 0.006 | -- | 0.000 | -- |
| **Hepatobiliary** |  |  |  |  |  |  |  |  |  |  |  |  |  |  |  |
| Simply adjusted model | 0.86 (0.83-0.89) | 858 | 1.00 (ref) | 695 | 0.85 (0.77-0.94) | 597 | 0.77 (0.69-0.85) | 560 | 0.75 (0.67-0.84) | 465 | 0.65 (0.58-0.73) | < 0.001 | < 0.001 | < 0.001 | < 0.001 |
| Main model | 0.90 (0.86-0.94) | 661 | 1.00 (ref) | 553 | 0.92 (0.82-1.03) | 464 | 0.83 (0.74-0.94) | 437 | 0.84 (0.74-0.95) | 367 | 0.75 (0.65-0.86) | < 0.001 | < 0.001 | < 0.001 | < 0.001 |
| **Pancreas** |  |  |  |  |  |  |  |  |  |  |  |  |  |  |  |
| Simply adjusted model | 0.88 (0.85-0.91) | 854 | 1.00 (ref) | 721 | 0.88 (0.80-0.97) | 621 | 0.80 (0.72-0.88) | 597 | 0.80 (0.72-0.89) | 493 | 0.69 (0.62-0.77) | < 0.001 | < 0.001 | < 0.001 | < 0.001 |
| Main model | 0.92 (0.88-0.96) | 656 | 1.00 (ref) | 577 | 0.96 (0.85-1.07) | 481 | 0.85 (0.75-0.96) | 468 | 0.89 (0.78-1.00) | 394 | 0.79 (0.69-0.91) | 0.005 | 0.009 | < 0.001 | < 0.001 |
| Additionally adjusted^c^ | 0.93  (0.89-0.97) | 656 | 1.00 (ref) | 577 | 0.97  (0.86-1.08) | 481 | 0.86  (0.77-0.98) | 468 | 0.90  (0.80-1.02) | 394 | 0.8  (0.70-0.92) | 0.011 | -- | 0.001 | -- |
| **Lung** |  |  |  |  |  |  |  |  |  |  |  |  |  |  |  |
| Simply adjusted model | 0.82 (0.80-0.84) | 1,375 | 1.00 (ref) | 1,073 | 0.82 (0.75-0.88) | 876 | 0.70 (0.64-0.76) | 769 | 0.65 (0.59-0.71) | 648 | 0.57 (0.52-0.63) | < 0.001 | < 0.001 | < 0.001 | < 0.001 |
| Main model | 0.88 (0.85-0.91) | 1,038 | 1.00 (ref) | 837 | 0.90 (0.82-0.98) | 663 | 0.77 (0.70-0.85) | 598 | 0.75 (0.68-0.83) | 506 | 0.68 (0.61-0.76) | < 0.001 | < 0.001 | < 0.001 | < 0.001 |
| Additionally adjusted^d^ | 0.88  (0.85-0.91) | 1,001 | 1.00 (ref) | 808 | 0.90  (0.82-0.98) | 643 | 0.77  (0.70-0.86) | 581 | 0.75  (0.68-0.84) | 491 | 0.68  (0.61-0.77) | < 0.001 | -- | < 0.001 | -- |
| **Malignant melanoma** |  |  |  |  |  |  |  |  |  |  |  |  |  |  |  |
| Simply adjusted model | 0.91 (0.88-0.94) | 966 | 1.00 (ref) | 864 | 0.92 (0.84-1.01) | 788 | 0.87 (0.79-0.96) | 762 | 0.87 (0.79-0.96) | 636 | 0.75 (0.68-0.83) | < 0.001 | < 0.001 | < 0.001 | < 0.001 |
| Main model | 0.92 (0.89-0.96) | 756 | 1.00 (ref) | 680 | 0.94 (0.85-1.05) | 622 | 0.90 (0.81-1.00) | 607 | 0.92 (0.82-1.03) | 507 | 0.80 (0.71-0.90) | 0.006 | 0.009 | 0.001 | 0.001 |
| Additionally adjusted^e^ | 0.93  (0.89-0.96) | 756 | 1.00 (ref) | 680 | 0.95  (0.85-1.05) | 622 | 0.90  (0.81-1.01) | 607 | 0.92  (0.83-1.03) | 507 | 0.80  (0.71-0.91) | 0.009 | -- | 0.001 | -- |
| **Connective soft tissue** |  |  |  |  |  |  |  |  |  |  |  |  |  |  |  |
| Simply adjusted model | 0.89 (0.85-0.92) | 717 | 1.00 (ref) | 627 | 0.91 (0.82-1.01) | 525 | 0.80 (0.71-0.89) | 501 | 0.79 (0.70-0.89) | 431 | 0.71 (0.63-0.80) | < 0.001 | < 0.001 | < 0.001 | < 0.001 |
| Main model | 0.91 (0.87-0.96) | 556 | 1.00 (ref) | 495 | 0.96 (0.85-1.08) | 407 | 0.84 (0.73-0.95) | 392 | 0.85 (0.75-0.98) | 338 | 0.77 (0.67-0.89) | 0.002 | 0.004 | < 0.001 | < 0.001 |
| **Breast** |  |  |  |  |  |  |  |  |  |  |  |  |  |  |  |
| Simply adjusted model | 0.93 (0.91-0.95) | 1,412 | 1.00 (ref) | 1,507 | 0.95 (0.88-1.02) | 1,476 | 0.89 (0.83-0.96) | 1,431 | 0.85 (0.79-0.91) | 1,355 | 0.81 (0.75-0.87) | < 0.001 | < 0.001 | < 0.001 | < 0.001 |
| Main model | 0.96 (0.93-0.99) | 1,067 | 1.00 (ref) | 1,163 | 0.99 (0.91-1.07) | 1,131 | 0.93 (0.85-1.01) | 1,112 | 0.91 (0.83-0.99) | 1,072 | 0.89 (0.81-0.97) | 0.048 | 0.051 | 0.003 | 0.003 |
| Additionally adjusted^f^ | 0.99  (0.95-1.03) | 539 | 1.00 (ref) | 756 | 1.06  (0.94-1.18) | 812 | 1.02  (0.91-1.13) | 815 | 0.97  (0.87-1.09) | 825 | 0.97  (0.86-1.08) | 0.434 | -- | 0.226 | -- |
| **Uterus** |  |  |  |  |  |  |  |  |  |  |  |  |  |  |  |
| Simply adjusted model | 0.87 (0.84-0.90) | 847 | 1.00 (ref) | 759 | 0.89 (0.80-0.98) | 668 | 0.80 (0.72-0.88) | 640 | 0.78 (0.70-0.87) | 531 | 0.67 (0.60-0.75) | < 0.001 | < 0.001 | < 0.001 | < 0.001 |
| Main model | 0.93 (0.90-0.97) | 645 | 1.00 (ref) | 597 | 0.98 (0.87-1.09) | 514 | 0.89 (0.79-1.00) | 501 | 0.92 (0.82-1.04) | 415 | 0.82 (0.71-0.93) | 0.020 | 0.024 | 0.002 | 0.002 |
| Additionally adjusted^g^ | 0.95  (0.89-1.01) | 229 | 1.00 (ref) | 300 | 1.07  (0.90-1.26) | 297 | 1.00  (0.84-1.19) | 290 | 1.01  (0.85-1.21) | 241 | 0.85  (0.70-1.03) | 0.127 | -- | 0.681 | -- |
| **Ovary** |  |  |  |  |  |  |  |  |  |  |  |  |  |  |  |
| Simply adjusted model | 0.90 (0.86-0.93) | 772 | 1.00 (ref) | 701 | 0.91 (0.82-1.01) | 612 | 0.82 (0.74-0.91) | 586 | 0.81 (0.72-0.90) | 515 | 0.73 (0.66-0.82) | < 0.001 | < 0.001 | < 0.001 | < 0.001 |
| Main model | 0.92 (0.88-0.96) | 595 | 1.00 (ref) | 549 | 0.96 (0.85-1.08) | 466 | 0.85 (0.75-0.96) | 456 | 0.87 (0.77-0.99) | 402 | 0.80 (0.70-0.92) | 0.008 | 0.010 | 0.001 | 0.001 |
| Additionally adjusted^g^ | 0.96  (0.89-1.02) | 175 | 1.00 (ref) | 247 | 1.11  (0.91-1.34) | 248 | 1.04  (0.86-1.27) | 244 | 1.00  (0.82-1.22) | 222 | 0.91  (0.74-1.12) | 0.324 | -- | 0.198 | -- |
| **Prostate** |  |  |  |  |  |  |  |  |  |  |  |  |  |  |  |
| Simply adjusted model | 0.98 (0.96-1.00) | 2,107 | 1.00 (ref) | 1,852 | 1.04 (0.98-1.11) | 1,570 | 1.00 (0.94-1.07) | 1,452 | 1.01 (0.94-1.08) | 1,290 | 0.95 (0.89-1.02) | 0.209 | 0.209 | 0.208 | 0.208 |
| Main model | 0.98 (0.95-1.00) | 1,717 | 1.00 (ref) | 1,528 | 1.05 (0.98-1.12) | 1,296 | 0.99 (0.92-1.07) | 1,181 | 1.00 (0.92-1.08) | 1,056 | 0.94 (0.87-1.03) | 0.171 | 0.171 | 0.163 | 0.163 |
| Additionally adjusted^h^ | 0.98  (0.96-1.01) | 1,672 | 1.00 (ref) | 1,488 | 1.05  (0.97-1.12) | 1,269 | 1.00  (0.92-1.07) | 1,153 | 1.00  (0.92-1.08) | 1,035 | 0.95  (0.87-1.03) | 0.217 | -- | 0.173 | -- |
| **Kidney** |  |  |  |  |  |  |  |  |  |  |  |  |  |  |  |
| Simply adjusted model | 0.86 (0.84-0.89) | 895 | 1.00 (ref) | 767 | 0.90 (0.82-0.99) | 627 | 0.77 (0.70-0.86) | 595 | 0.76 (0.69-0.85) | 492 | 0.66 (0.59-0.73) | < 0.001 | < 0.001 | < 0.001 | < 0.001 |
| Main model | 0.90 (0.87-0.94) | 694 | 1.00 (ref) | 599 | 0.95 (0.85-1.06) | 489 | 0.83 (0.74-0.93) | 466 | 0.85 (0.75-0.96) | 387 | 0.75 (0.65-0.85) | < 0.001 | < 0.001 | < 0.001 | < 0.001 |
| **Bladder** |  |  |  |  |  |  |  |  |  |  |  |  |  |  |  |
| Simply adjusted model | 0.88 (0.85-0.91) | 863 | 1.00 (ref) | 728 | 0.90 (0.81-0.99) | 603 | 0.79 (0.71-0.88) | 581 | 0.80 (0.72-0.89) | 488 | 0.70 (0.63-0.79) | < 0.001 | < 0.001 | < 0.001 | < 0.001 |
| Main model | 0.92 (0.88-0.96) | 670 | 1.00 (ref) | 576 | 0.95 (0.85-1.07) | 473 | 0.85 (0.75-0.95) | 464 | 0.89 (0.79-1.01) | 389 | 0.80 (0.70-0.91) | 0.006 | 0.009 | 0.001 | 0.001 |
| **Brain** |  |  |  |  |  |  |  |  |  |  |  |  |  |  |  |
| Simply adjusted model | 0.90 (0.86-0.93) | 781 | 1.00 (ref) | 683 | 0.91 (0.82-1.01) | 576 | 0.80 (0.72-0.90) | 555 | 0.81 (0.72-0.90) | 480 | 0.72 (0.64-0.81) | < 0.001 | < 0.001 | < 0.001 | < 0.001 |
| Main model | 0.92 (0.88-0.96) | 608 | 1.00 (ref) | 542 | 0.96 (0.85-1.08) | 440 | 0.82 (0.72-0.93) | 433 | 0.86 (0.75-0.97) | 380 | 0.79 (0.68-0.90) | 0.001 | 0.002 | < 0.001 | < 0.001 |
| **Thyroid** |  |  |  |  |  |  |  |  |  |  |  |  |  |  |  |
| Simply adjusted model | 0.89 (0.85-0.92) | 741 | 1.00 (ref) | 638 | 0.89 (0.80-0.99) | 536 | 0.77 (0.69-0.87) | 535 | 0.80 (0.71-0.89) | 454 | 0.70 (0.62-0.79) | < 0.001 | < 0.001 | < 0.001 | < 0.001 |
| Main model | 0.91 (0.87-0.95) | 574 | 1.00 (ref) | 504 | 0.94 (0.83-1.06) | 414 | 0.81 (0.71-0.92) | 421 | 0.87 (0.76-0.99) | 352 | 0.76 (0.66-0.87) | 0.001 | 0.002 | < 0.001 | < 0.001 |
| **Lymphoid and Hematopoietic Tissues** |  |  |  |  |  |  |  |  |  |  |  |  |  |  |  |
| Simply adjusted model | 0.91 (0.88-0.93) | 1,239 | 1.00 (ref) | 1,077 | 0.91 (0.84-0.99) | 936 | 0.84 (0.77-0.91) | 900 | 0.84 (0.77-0.92) | 776 | 0.76 (0.69-0.83) | < 0.001 | < 0.001 | < 0.001 | < 0.001 |
| Main model | 0.93 (0.90-0.97) | 961 | 1.00 (ref) | 841 | 0.95 (0.86-1.04) | 731 | 0.88 (0.79-0.97) | 700 | 0.90 (0.81-0.99) | 607 | 0.82 (0.74-0.91) | 0.004 | 0.008 | < 0.001 | < 0.001 |

Abbreviations: LA%, LA to total fatty acids percentage; SD, standard deviation; CI, confidence interval; HR, hazards ratio; ref, reference.

The results from simply adjusted models revealed the associations of plasma LA% with cancer risk stratified by age and sex in general cohort. The main models were adjusted for general covariates including ethnicity (classified into White, Black, Asian, Others), Townsend deprivation index (continuous), assessment Center, BMI (kg/m2; continuous), smoking status (categorized as never, previous, current), alcohol intake status (categorized as never, previous, current), and physical activity (classified as low, moderate, high). The additionally adjusted models were adjusted for extra covariates for some specific types of cancer.

^a^ Additionally adjusted for gastroesophageal reflux disease at baseline and waist-hip ratio.

^b^ Additionally adjusted for diabetes at baseline, aspirin use, processed meat intake, waist-hip ratio, and family history.

^c^ Additionally adjusted for diabetes at baseline.

^d^ Additionally adjusted for family history.

^e^ Additionally adjusted for skin color, ease of skin tanning, use of sun/UV protection, childhood sunburn occasions, frequency of solarium/sunlamp use.

^f^ Restricted to female, and additionally adjusted for age when menarche started, hormone replacement therapy use, oral contraceptive use, number of live births, menopausal status, hysterectomy status, and family history.

^g^ Restricted to female, and additionally adjusted for age when menarche started, hormone replacement therapy use, oral contraceptive use, number of live births, menopausal status, hysterectomy status.

^h^ Restricted to male, and additionally adjusted for family history.

^i^ Used likelihood ratio test to compare the full model with reduced model.

^j^ Based on False Discovery Rate (FDR) to calculate the adjusted p-values for simply adjusted models and main models among 19 cancer sites.

^k^ Used the median value of each quintile as a continuous variable within the models.

Table S8. Associations^a^ of plasma omega-6% and omega-3% with incidence of overall cancer in the UK Biobank Study, excluding those who had outcome in the first follow-up year

| Variable forms | **Overall Cancer** | | | | | | |
| --- | --- | --- | --- | --- | --- | --- | --- |
|  | Omega-6 | | |  | Omega-3 | | |
|  | Events | HR  (95% CI) | P values |  | Events | HR  (95% CI) | P values |
| Continuous (per 1-SD) | 22,021 | 0.98  (0.96-0.99) | 0.001 |  | 22,021 | 0.98  (0.97-1.00) | 0.027 |
| Quintiles |  |  | | | | | |
| 1 | 5,005 | 1.00  (ref) | -- |  | 4,253 | 1.00  (ref) | -- |
| 2 | 4,718 | 0.99  (0.95-1.03) | 0.541 |  | 4,301 | 0.97  (0.93-1.01) | 0.147 |
| 3 | 4,468 | 0.98  (0.94-1.02) | 0.356 |  | 4,367 | 0.95  (0.91-0.99) | 0.015 |
| 4 | 4,099 | 0.95  (0.91-0.99) | 0.015 |  | 4,486 | 0.95  (0.91-0.99) | 0.015 |
| 5 | 3,731 | 0.94  (0.90-0.98) | 0.007 |  | 4,614 | 0.95  (0.91-0.99) | 0.023 |
| *P for overall* | *0.029* | | |  | *0.076* | | |
| *P for trend* | *0.002* | | |  | *0.028* | | |

Abbreviations: omega-6%, omega-6 fatty acids to total fatty acids percentage; omega-3%, omega-3 fatty acids to total fatty acids percentage; CI, confidence interval; HR, hazards ratio; ref, reference.

^a^ From Cox proportional hazards regression; results were based on the main models, stratified by age and sex, and adjusted for ethnicity (classified into White, Black, Asian, Others), Townsend deprivation index (continuous), assessment Center, BMI (kg/m2; continuous), smoking status (categorized as never, previous, current), alcohol intake status (categorized as never, previous, current), and physical activity (classified as low, moderate, high).

Table S9. Associations^a^ of plasma omega-6% and omega-3% with incidence of overall cancer in the UK Biobank Study, excluding those who had outcome in the first-three follow-up years

| Variable forms | **Overall Cancer** | | | | | | |
| --- | --- | --- | --- | --- | --- | --- | --- |
|  | Omega-6 | | |  | Omega-3 | | |
|  | Events | HR  (95% CI) | P values |  | Events | HR  (95% CI) | P values |
| Continuous (per 1-SD) | 18,457 | 0.97  (0.96-0.99) | < 0.001 |  | 18,457 | 0.98  (0.97-1.00) | 0.039 |
| Quintiles |  |  | | | | | |
| 1 | 4,237 | 1.00  (ref) | -- |  | 3,577 | 1.00  (ref) | -- |
| 2 | 3,945 | 0.98  (0.94-1.02) | 0.352 |  | 3,625 | 0.97  (0.93-1.02) | 0.244 |
| 3 | 3,727 | 0.97  (0.93-1.02) | 0.219 |  | 3,674 | 0.95  (0.91-1.00) | 0.041 |
| 4 | 3,438 | 0.94  (0.90-0.99) | 0.016 |  | 3,734 | 0.95  (0.90-0.99) | 0.019 |
| 5 | 3,110 | 0.93  (0.88-0.97) | 0.002 |  | 3,847 | 0.95  (0.91-1.00) | 0.053 |
| *P for overall* | *0.024* | | |  | *0.140* | | |
| *P for trend* | *0.001* | | |  | *0.047* | | |

Abbreviations: omega-6%, omega-6 fatty acids to total fatty acids percentage; omega-3%, omega-3 fatty acids to total fatty acids percentage; CI, confidence interval; HR, hazards ratio; ref, reference.

^a^ From Cox proportional hazards regression; results were based on the main models, stratified by age and sex, and adjusted for ethnicity (classified into White, Black, Asian, Others), Townsend deprivation index (continuous), assessment Center, BMI (kg/m2; continuous), smoking status (categorized as never, previous, current), alcohol intake status (categorized as never, previous, current), and physical activity (classified as low, moderate, high).

Table S10. Baseline characteristics of participants with and without exposure information

| **Characteristics**^a^ | Participants missed PUFAs information  **(N=210,264)** | Participants had PUFAs information  **(N=253,138)** |
| --- | --- | --- |
| **Age** (years) | 56.3 (8.1) | 56.4 (8.1) |
| **Gender** (male%) | 97,324 (46.3%) | 119,450 (47.2%) |
| **Ethnicity**(n%) |  |  |
| White | 188,159 (90.1%) | 230,222 (91.4%) |
| Black | 1,368 (0.7%) | 1,377 (0.5%) |
| Asian | 8,751 (4.2%) | 8,979 (3.6%) |
| Others | 10,537 (5.0%) | 11,412 (4.5%) |
| Missing (n) | *1,449* | *1,148* |
| **TDI** | -1.2 (3.1) | -1.4 (3.1) |
| Missing (n) | 267 | 311 |
| **BMI** (kg/m2) | 27.4 (4.8) | 27.5 (4.8) |
| Missing (n) | *1,886* | *949* |
| **Smoking status** (n%) |  |  |
| Never | 115,429 (55.3%) | 138,605 (55.0%) |
| Previous | 71,117 (34.1%) | 86,723 (34.4%) |
| Current | 22,255 (10.7%) | 26,593 (10.6%) |
| Missing (n) | *1,463* | *1,217* |
| **Alcohol status** (n%) |  |  |
| Never | 9,656 (4.6%) | 10,969 (4.3%) |
| Previous | 7,520 (3.6%) | 8,897 (3.5%) |
| Current | 192,155 (91.8%) | 232,667 (92.1%) |
| Missing (n) | *933* | *605* |
| **Physical activity** (n%) |  |  |
| Low | 31,325 (18.7%) | 38,683 (18.9%) |
| Moderate | 68,713 (41.1%) | 82,534 (40.4%) |
| High | 67,216 (40.2%) | 83,303 (40.7%) |
| Missing (n) | *43,010* | *48,618* |

^a^ All variables measured at baseline are presented as mean (SD) except as otherwise specified.

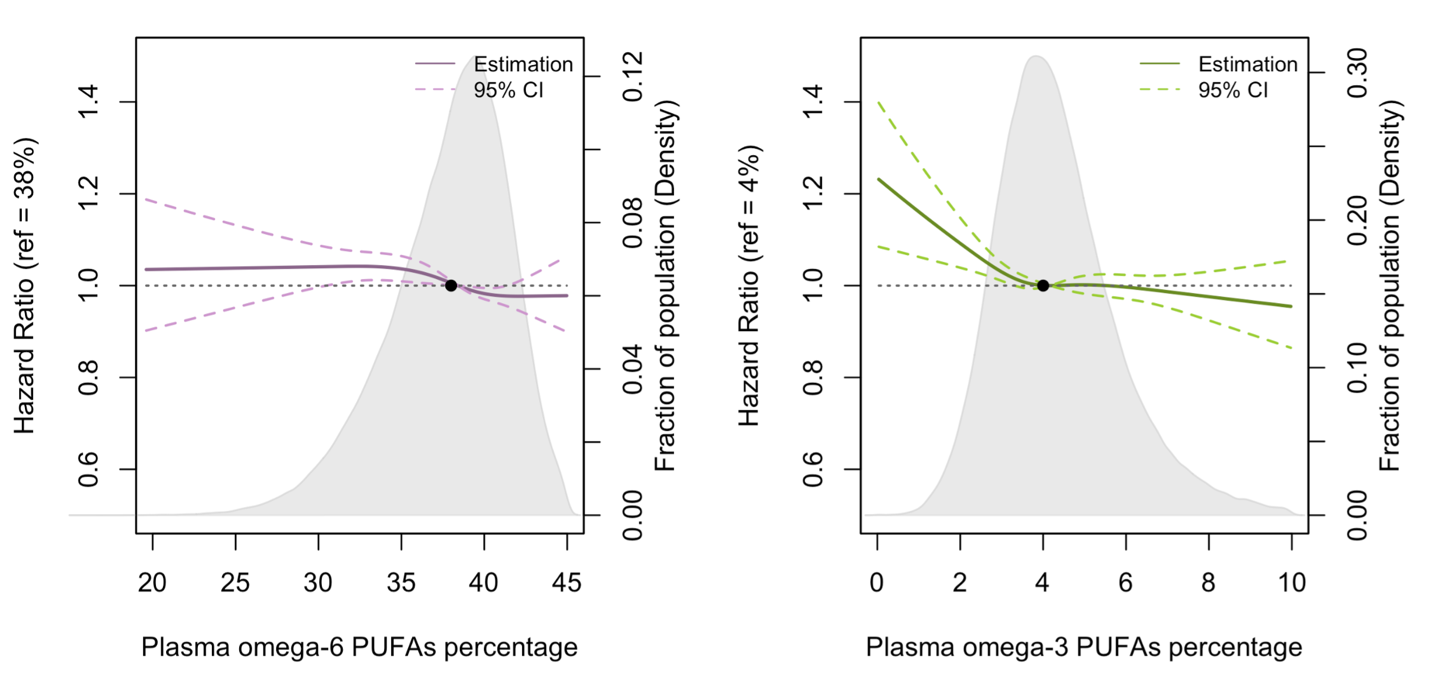

**Figure S2. Associations of plasma omega-6% and omega-3% with overall cancer incidence evaluated using restricted cubic splines. Hazard ratios and plasma PUFAs percentage are presented in the vertical and horizontal axis, respectively.** The best estimates and their confidence intervals are presented as solid lines and dotted lines, respectively. The median of the percentages (38% for omega-6 PUFAs and 4% for omega-3 PUFAs) were selected as the reference levels. Potential nonlinearity was identified for omega-3% with overall cancer (p < 0.05). All HRs are stratified by age and sex, and adjusted for ethnicity (classified into White, Black, Asian, Others), Townsend deprivation index (continuous), assessment Center, BMI (kg/m2; continuous), smoking status (categorized as never, previous, current), alcohol intake status (categorized as never, previous, current), and physical activity (classified as low, moderate, high).

**Figure S3. Associations of plasma omega-6% and omega-3% with prostate cancer incidence evaluated using restricted cubic splines. Hazard ratios and plasma PUFAs percentage are presented in the vertical and horizontal axis, respectively.** The best estimates and their confidence intervals are presented as solid lines and dotted lines, respectively. The median of the percentages (38% for omega-6 PUFAs and 4% for omega-3 PUFAs) were selected as the reference levels. Potential nonlinearity was identified for omega-6% with prostate cancer (p = 0.02). All HRs are stratified by age and sex, and adjusted for ethnicity (classified into White, Black, Asian, Others), Townsend deprivation index (continuous), assessment Center, BMI (kg/m2; continuous), smoking status (categorized as never, previous, current), alcohol intake status (categorized as never, previous, current), and physical activity (classified as low, moderate, high).

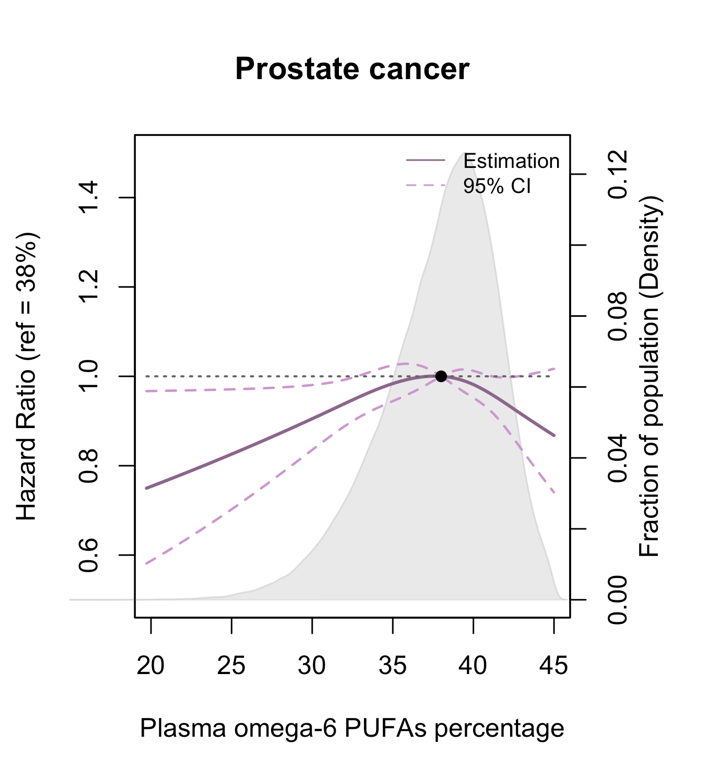

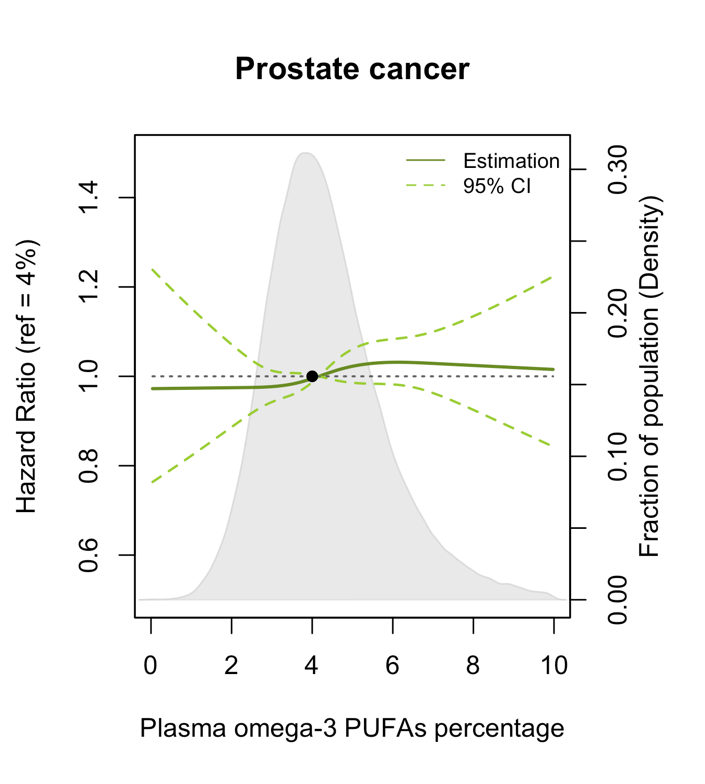

Table S11. STROBE Statement—Checklist of items that should be included in reports of ***cohort studies***

|  | Item No | Recommendation | Page No |
| --- | --- | --- | --- |
| **Title and abstract** | 1 | (*a*) Indicate the study’s design with a commonly used term in the title or the abstract |  |
|  |  | (*b*) Provide in the abstract an informative and balanced summary of what was done and what was found | 1-2 |
| Introduction | | | |
| Background/rationale | 2 | Explain the scientific background and rationale for the investigation being reported | 3-4 |
| Objectives | 3 | State specific objectives, including any prespecified hypotheses | 4 |
| Methods | | | |
| Study design | 4 | Present key elements of study design early in the paper | 4 |
| Setting | 5 | Describe the setting, locations, and relevant dates, including periods of recruitment, exposure, follow-up, and data collection | 4 |
| Participants | 6 | (*a*) Give the eligibility criteria, and the sources and methods of selection of participants. Describe methods of follow-up | 4 |
|  |  | (*b*) For matched studies, give matching criteria and number of exposed and unexposed |  |
| Variables | 7 | Clearly define all outcomes, exposures, predictors, potential confounders, and effect modifiers. Give diagnostic criteria, if applicable | 4-5 |
| Data sources/ measurement | 8* | For each variable of interest, give sources of data and details of methods of assessment (measurement). Describe comparability of assessment methods if there is more than one group | 4-5 |
| Bias | 9 | Describe any efforts to address potential sources of bias | 4 |
| Study size | 10 | Explain how the study size was arrived at | 4 |
| Quantitative variables | 11 | Explain how quantitative variables were handled in the analyses. If applicable, describe which groupings were chosen and why | 4-5 |
| Statistical methods | 12 | (*a*) Describe all statistical methods, including those used to control for confounding |  |
|  |  | (*b*) Describe any methods used to examine subgroups and interactions |  |
|  |  | (*c*) Explain how missing data were addressed | 6-7 |
|  |  | (*d*) If applicable, explain how loss to follow-up was addressed |  |
|  |  | (*e*) Describe any sensitivity analyses |  |
| Results | | |  |
| Participants | 13* | (a) Report numbers of individuals at each stage of study—eg numbers potentially eligible, examined for eligibility, confirmed eligible, included in the study, completing follow-up, and analysed | 8 |
|  |  | (b) Give reasons for non-participation at each stage |  |
|  |  | (c) Consider use of a flow diagram |  |
| Descriptive data | 14* | (a) Give characteristics of study participants (eg demographic, clinical, social) and information on exposures and potential confounders | 8 |
|  |  | (b) Indicate number of participants with missing data for each variable of interest |  |
|  |  | (c) Summarise follow-up time (eg, average and total amount) |  |
| Outcome data | 15* | Report numbers of outcome events or summary measures over time | 8 |

| Main results | 16 | (*a*) Give unadjusted estimates and, if applicable, confounder-adjusted estimates and their precision (eg, 95% confidence interval). Make clear which confounders were adjusted for and why they were included | 8-10 |
| --- | --- | --- | --- |
|  |  | (*b*) Report category boundaries when continuous variables were categorized |  |
|  |  | (*c*) If relevant, consider translating estimates of relative risk into absolute risk for a meaningful time period |  |
| Other analyses | 17 | Report other analyses done—eg analyses of subgroups and interactions, and sensitivity analyses | 10-11 |
| Discussion | | | |
| Key results | 18 | Summarise key results with reference to study objectives | 11 |
| Limitations | 19 | Discuss limitations of the study, taking into account sources of potential bias or imprecision. Discuss both direction and magnitude of any potential bias | 14-15 |
| Interpretation | 20 | Give a cautious overall interpretation of results considering objectives, limitations, multiplicity of analyses, results from similar studies, and other relevant evidence | 11-14 |
| Generalisability | 21 | Discuss the generalisability (external validity) of the study results | 15 |
| Other information | | | |
| Funding | 22 | Give the source of funding and the role of the funders for the present study and, if applicable, for the original study on which the present article is based | 17 |

*Give information separately for exposed and unexposed groups.

**Note:** An Explanation and Elaboration article discusses each checklist item and gives methodological background and published examples of transparent reporting. The STROBE checklist is best used in conjunction with this article (freely available on the Web sites of PLoS Medicine at http://www.plosmedicine.org/, Annals of Internal Medicine at http://www.annals.org/, and Epidemiology at http://www.epidem.com/). Information on the STROBE Initiative is available at http://www.strobe-statement.org.
